# The Alzheimer’s Association Global Biomarker Standardization Consortium (GBSC) plasma phospho-tau Round Robin study

**DOI:** 10.1101/2024.08.22.24312244

**Authors:** Nicholas J. Ashton, Ashvini Keshavan, Wagner S. Brum, Ulf Andreasson, Burak Arslan, Mathias Droescher, Stefan Barghorn, Jeroen Vanbrabant, Charlotte Lambrechts, Maxime Van Loo, Erik Stoops, Shweta Iyengar, HaYeun Ji, Xiaomei Xu, Alex Forrest-Hay, Bingqing Zhang, Yuling Luo, Andreas Jeromin, Manu Vandijck, Nathalie Le Bastard, Hartmuth Kolb, Gallen Triana-Baltzer, Divya Bali, Shorena Janelidze, Shieh-Yueh Yang, Catherine Demos, Daniel Romero, George Sigal, Jacob Wohlstadter, Kishore Malyavantham, Meenakshi Khare, Alexander Jethwa, Laura Stoeckl, Johan Gobom, Przemysław R. Kac, Fernando Gonzalez-Ortiz, Laia Montoliu-Gaya, Oskar Hansson, Robert A. Rissman, Maria C. Carillo, Leslie M Shaw, Kaj Blennow, Jonathan M. Schott, Henrik Zetterberg

## Abstract

**BACKGROUND:** Phosphorylated tau (p-tau) is a specific blood biomarker for Alzheimer’s disease (AD) pathology. Multiple p-tau biomarkers on several analytical platforms are poised for clinical use. The Alzheimer’s Association Global Biomarker Standardisation Consortium plasma phospho-tau Round Robin study engaged assay developers in a blinded case-control study on plasma p-tau, aiming to learn which assays provide the largest fold-changes in AD compared to non-AD, have the strongest relationship between plasma and cerebrospinal fluid (CSF), and show the most consistent relationships between methods (commutability) in measuring both patient samples and candidate reference materials (CRM).

**METHODS:** Thirty-three different p-tau biomarker assays, built on eight different analytical platforms, were used to quantify paired plasma and CSF samples from 40 participants. AD biomarker status was categorised as “AD pathology” (n=25) and “non-AD pathology” (n=15) by CSF Aβ42/Aβ40 (US-FDA; CE-IVDR) and p-tau181 (CE-IVDR) methods. The commutability of four CRM, at three concentrations, was assessed across assays.

**FINDINGS:** Plasma p-tau217 consistently demonstrated higher fold-changes between AD and non-AD pathology groups, compared to other p-tau epitopes. Fujirebio LUMIPULSE G, UGOT IPMS, and Lilly MSD p-tau217 assays provided the highest median fold-changes. In CSF, p-tau217 assays also performed best, and exhibited substantially larger fold-changes than their plasma counterparts, despite similar diagnostic performance. P-tau217 showed the strongest correlations between plasma assays (rho=0.81 to 0.97). Plasma p-tau levels were weakly-to-moderately correlated with CSF p-tau, and correlations were non-significant within the AD group alone. The evaluated CRM were not commutable across assays.

**INTERPRETATION:** Plasma p-tau217 measures had larger fold-changes and discriminative accuracies for detecting AD pathology, and better agreement across platforms than other plasma p-tau variants. Plasma and CSF markers of p-tau, measured by immunoassays, are not substantially correlated, questioning the interchangeability of their continuous relationship. Further work is warranted to understand the pathophysiology underlying this dissociation, and to develop suitable reference materials facilitating cross-assay standardisation.

**FUNDING:** Alzheimer’s Association (#ADSF-24-1284328-C)

**RESEARCH IN CONTEXT:** *Evidence before this study:* Phosphorylated tau (p-tau) in cerebrospinal fluid is an established biomarker of Alzheimer’s disease (AD), and several studies of plasma phosphorylated tau (p-tau) biomarkers now show evidence for its diagnostic accuracy in AD. We reviewed the literature on plasma p-tau in AD by searching without language restrictions in PubMed for articles from 1^st^ January 2014 to 1^st^ March 2024 using the terms ((("Alzheimer Disease"[Mesh]) OR ((Alzheimer[Title/Abstract]) OR (Alzheimer’s [Title/Abstract))) AND ((("Blood"[Mesh]) OR ("Plasma"[Mesh])) OR (plasma[Title/Abstract]))) AND ((phospho tau[Title/Abstract]) OR (phosphorylated tau[Title/Abstract]) OR (p-tau[Title/Abstract]) OR (P-tau[Title/Abstract]) OR (pTau[Title/Abstract])), filtering for studies in humans. Several studies examined plasma p-tau181, p-tau217 and p-tau231 in well-characterised research cohorts including cognitively impaired and unimpaired individuals, demonstrating high diagnostic accuracy when utilising CSF or amyloid PET imaging “gold standard” biomarkers, and in relation to post-mortem amyloid and tau pathology. Most studies utilised single-method assays to measure biomarkers of interest, including immunoprecipitation-mass spectrometry and a variety of immunochemical methods including single molecule array, enzyme-linked immunosorbent assays, electrochemiluminescence, and immunomagnetic reduction. Some later studies have performed cross-method and cross-phospho-form comparisons (p-tau181, p-tau217 and p-tau231), including six single molecular array assays in the Amsterdam Dementia Cohort, nine assays in the BIODEGMAR memory clinic-based cohort from Spain, ten assays in individuals with mild cognitive impairment in the Skåne University memory clinic cohort from Sweden, and have generally reported the various individual methods for p-tau217 to have the highest diagnostic accuracy for the presence of cerebral β-amyloid deposition. No study to date has compared assays for p-tau217 on semi- and fully automated platforms, described comparisons with p-tau212 and p-tau205 in plasma, or assessed commutability of candidate reference materials between assays in comparison to patient samples.

*Added value of this study:* Our study included the largest number of methods for assaying plasma p-tau to date, with engagement from 12 assay developers, allowing for blinded cross-assay comparisons of 33 plasma assays (for p-tau181, p-tau205, p-tau212, p-tau217, and p-tau231). Among these were included several semi-automated and fully automated methods that have potential for widespread clinical application. This is also the first study, to our knowledge, in which the commutability of candidate reference materials was assessed for plasma p-tau, as a first step in efforts to standardise between assays. The study design capitalised on a large volume of paired CSF and plasma drawn from well-characterised attenders to a specialist cognitive disorders service, allowing for 26 intra-method comparisons between CSF and plasma measurements within the same individuals. We found that CSF p-tau217 assays consistently outperformed other CSF p-tau assays in terms of fold-change between individuals with and without Alzheimer’s pathology, and the top ten assays in terms of fold change in plasma were also p-tau217 assays. This high discriminative value of plasma p-tau217 contrasted with overall only moderate correlations between CSF and plasma within individuals, which did not persist within the AD group alone. We also did not observe commutability of the four types of candidate reference materials we investigated, for any of the assay pairs.

*Implications of all the available evidence:* Our study adds to the growing evidence for plasma p-tau217 as a candidate biomarker for translation into clinical practice by virtue of its superior discriminative ability between AD vs non-AD in symptomatic individuals, when compared head-to-head with other phosphorylated tau forms. The observed high and clinically interpretable fold-changes are pivotal for this biomarker’s possible future success. Further studies should examine whether blood biomarker-supported diagnosis will extend access to disease-specific (and potentially disease-modifying) treatments and impact patient-relevant outcomes such as quality of life, particularly in diverse and resource-limited settings. Standardization between different assays will also be important for real-world applications, and this will require further concerted efforts in developing reference materials.

## INTRODUCTION

The neuropathological confirmation of amyloid-β (Aβ) plaques and tau neurofibrillary tangles (NFT) remains the gold standard for a definitive diagnosis of Alzheimer’s disease (AD). However, the clinical assessment of AD is being increasingly supported by validated positron emission tomography (PET) imaging and cerebrospinal fluid (CSF) biomarkers accurately reflecting Aβ “A”, tau “T”, and neurodegeneration “N” pathologies, which have improved the accuracy in diagnosing AD during life, and provided evidence for a biological classification of the disease (AT(N)).^1^ Yet, such biomarkers are considered to be specialised and have significant constraints (eg, invasiveness and skill for CSF sampling, and cost for PET imaging) hindering their use as general tools for diagnosing and managing dementia in health systems across the globe.

Blood biomarkers capable of detecting core AD pathologies have demonstrated huge potential for clinical practice use, and in determining eligibility for, and response to, novel treatments. ^2,3^ Plasma amyloid-β peptides (Aβ42/Aβ40),^4–6^ neurofilament light (NfL)^7,8^ and glial fibrillary acidic protein (GFAP)^9,10^ have all been shown to associate with certain AD features, but none can demonstrate the high disease-specificity of plasma phosphorylated tau (p-tau). Increased p-tau is initially associated with Aβ deposition in asymptomatic individuals; further increases are seen in the symptomatic phases of AD, when overt NFT pathology is present in the brain and driving cognitive symptoms.

Tau is present and detectable in blood in various phosphorylated forms, including (but not limited to) p-tau181, p-tau205, p-tau212, p-tau217, and p-tau231. Non-phosphorylated tau is also detectable in blood, either as “total-tau”,^11^ “brain-derived” tau,^12^ “N-terminal tau [NTA]” tau,^13^ or as non-phosphorylated peptide forms.^14^ Despite significant changes in symptomatic disease, non-phosphorylated tau species have limited utility in AD diagnostics but have possible application in acute neurological conditions^15^ or more advanced disease stages.^13^ For the most part, p-tau epitopes in blood exhibit a similar pattern of increase as AD pathology develops. However, distinctions have been reported between p-tau forms in terms of diagnostic accuracy in symptomatic individuals,^16–18^ relationships with in vivo and post-mortem pathology,^19,20^ preclinical detection,^21,22^ physiological fluctuations,^23^ and longitudinal change.^21^ These findings suggest that constructing disease staging based on biofluid measures of tau may be feasible.^24^

In addition to any possible pathophysiological differences between phospho-forms, different quantification methods may also differentially influence results. Since the initial studies piloting p-tau detection in blood,^25,26^ several variations of antibody-based technologies have been developed for quantification at femtomolar concentrations (eg, Single molecule array [Simoa],^27,28^ immunomagnetic reduction [IMR],^29^ electrochemiluminescence [eg, MesoScale Discovery;^30^ Elecsys^®^, Roche Diagnostics International Ltd, Rotkreuz, Switzerland],^31^ and immunoprecipitation mass spectrometry [IPMS]^14,32^). As recently approved anti-Aβ therapies for AD approach clinical implementation, use of numerous validated measures of blood p-tau will likely guide timely treatment decisions. Several studies have already compared different p-tau immunoassay platforms to detect a binary categorisation of AD pathology.^16,17,33–35^ Yet, it is also important to understand the translatability of different plasma p-tau results across multiple platforms.

In this Alzheimer’s Association Global Biomarker Standardisation Consortium plasma p-tau Round Robin study, we performed a comprehensive and blinded comparison of 33 different p-tau assays, including seven different p-tau epitopes, or p-tau/t-tau ratios, utilising eight immunological platforms, in plasma and CSF from symptomatic individuals categorised as having AD or non-AD pathology. Our main aim was to compare all assays regarding their ability to detect AD pathology (focusing on fold-change between AD and non-AD groups), correlations between plasma biomarkers and assays, and relationships with CSF p-tau. A secondary aim was to test the commutability of four candidate reference materials (CRM), ie, the consistency of relationships between assays in measuring the CRM compared with the participant samples.

## METHODS

### Participants, ethics, and study design

Individual de[identified EDTA plasma and CSF samples were from the prospective Wolfson CSF study 12/0344 (PI Schott; NRES London Queen Square August 2013) at the University College London Dementia Research Centre. All individuals were being investigated by lumbar puncture for cognitive concerns after having been assessed in the specialist cognitive disorders service at the National Hospital for Neurology and Neurosurgery, University College London Hospitals NHS Trust, London, UK. Participants gave informed written consent to opportunistic research sample donation at the same time as sampling of their CSF and paired venous plasma for diagnostic purposes. Participant samples were collected serially over the period December 2020 to June 2022, and selected based on known CSF Aβ42/Aβ40 and spanning a range of p-tau181 (LUMIPULSE G) concentrations previously measured in clinical routine, and availability of sufficient bio-banked CSF (total 4 mL) and plasma (total 7 mL). These total volumes determined after surveying all prospective participating labs to ascertain their minimal and ideal volumes of CSF and plasma required to carry out their respective assays. A participant was considered to have “AD pathology” if the CSF results were Aβ42/Aβ40 <0·065 and p-tau181 >57 pg/mL. Plasma and CSF aliquots of 1mL were sent on dry ice to the University of Gothenburg for sub-aliquoting and distribution to participating laboratories/assay developers, blinded to participant information.

### CSF and plasma collection

Participants were not instructed to fast, and CSF sampling was performed between 0800 and 1200 hours. After local anesthesia with lignocaine, a 22-gauge atraumatic spinal needle was used to collect up to 20 mL of CSF, without active withdrawal, into 2 × 10 mL polypropylene screw top containers (Sarstedt 62·610·018), which were transported at ambient temperature within 30 minutes to the laboratory. CSF was centrifuged at 1750 g for 5 minutes at 4°C and the supernatant placed in 1 mL aliquots into polypropylene screw top cryovials. Peripheral venous blood was sampled using a tourniquet and 21-gauge or 23-gauge butterfly needle with a BD Vacutainer collecting system, into 6 mL K3-EDTA plasma tubes, which were transported and centrifuged at ambient temperature, at 1800 g for 5 mins, within 30 mins of sampling. Plasma supernatant aliquots of 1 mL were stored in polypropylene screw-top cryovials. Both CSF and plasma were stored at −80°C within 60 minutes of sampling.

### Phosphorylated tau assays

Eleven participating centres received plasma and CSF aliquots. In total, 31 single p-tau measurements (eleven p-tau181, one p-tau205, one p-tau212, thirteen p-tau217, and five p-tau231) across eight immunological platforms were compared. In addition, we also included two p-tau/tau ratios derived from mass spectrometric measurements : p-tau205/tau205 (ie, p-tau205/tau195-209) and p-tau217/tau217 (ie, p-tau217/tau212-221). All measurements were made in duplicate, except for those undertaken on the fully automated instruments (LUMIPULSE G, Fujrebio Europe N.V., Ghent, Belgium and Cobas^®^ e 801 analyzer, Roche Diagnostics International Ltd, Rotkreuz, Switzerland) and NULISA^TM^. Each assay was performed in plasma and CSF except for the Elecsys pTau217 prototype immunoassay (Elecsys p-tau217; Roche Diagnostics International Ltd, Rotkreuz, Switzerland) and the

UGOT IPMS, which were not available for CSF. An overview of the immunological platforms is shown in Table 1 (Supplementary Table 1, appendix p18, if assay procedures differ for CSF). Methods have previously been described for ADx Simoa p-tau181,^33^ ALZpath p-tau217,^36^ Janssen Simoa p-tau217,^28,37^ Fujirebio Lumipulse G pTau181 (Plasma),^17^ MSD Lilly p-tau181 and p-tau217,^30,38,39^ MagQu p-tau181,^29^ Meso Scale S-PLEX p-tau181 ^40^ and p-tau217^41^, Quanterix Simoa p-tau181 v2.1,^42^ Roche Elecsys p-tau181 (Roche Diagnostics International Ltd, Rotkreuz, Switzerland),^31^ UGOT p-tau181,^27,43^ UGOT p-tau212,^44^ UGOT p-tau217,^45^ UGOT p-tau231,^46,47^ and UGOT IPMS.^14^ Method descriptions for Abbvie Erenna p-tau217 and p-tau231, ADx Lumipulse G p-tau217, ADx Simoa p-tau217, Alamar Biosciences NULISA p-tau181, p-tau217 and p-tau231, Fujirebio Lumipulse G pTau217 Plasma RUO, Meso Scale S-PLEX p-tau231, and Roche Elecsys p-tau217 are detailed in the Supplementary Methods (appendix p3-4). All assay measures were performed by assay vendors. CSF and plasma ALZpath p-tau217 were measured at the Department of Neurochemistry, University of Gothenburg. CSF and plasma MSD Lilly p-tau217 and p-tau181 were measured at the Clinical Memory Research Unit, Lund University. Analytical performance of the assays in terms of repeatability, intermediate precision, and sample performance is shown in Supplementary Table 2 (appendix p19).

**Table 1.**
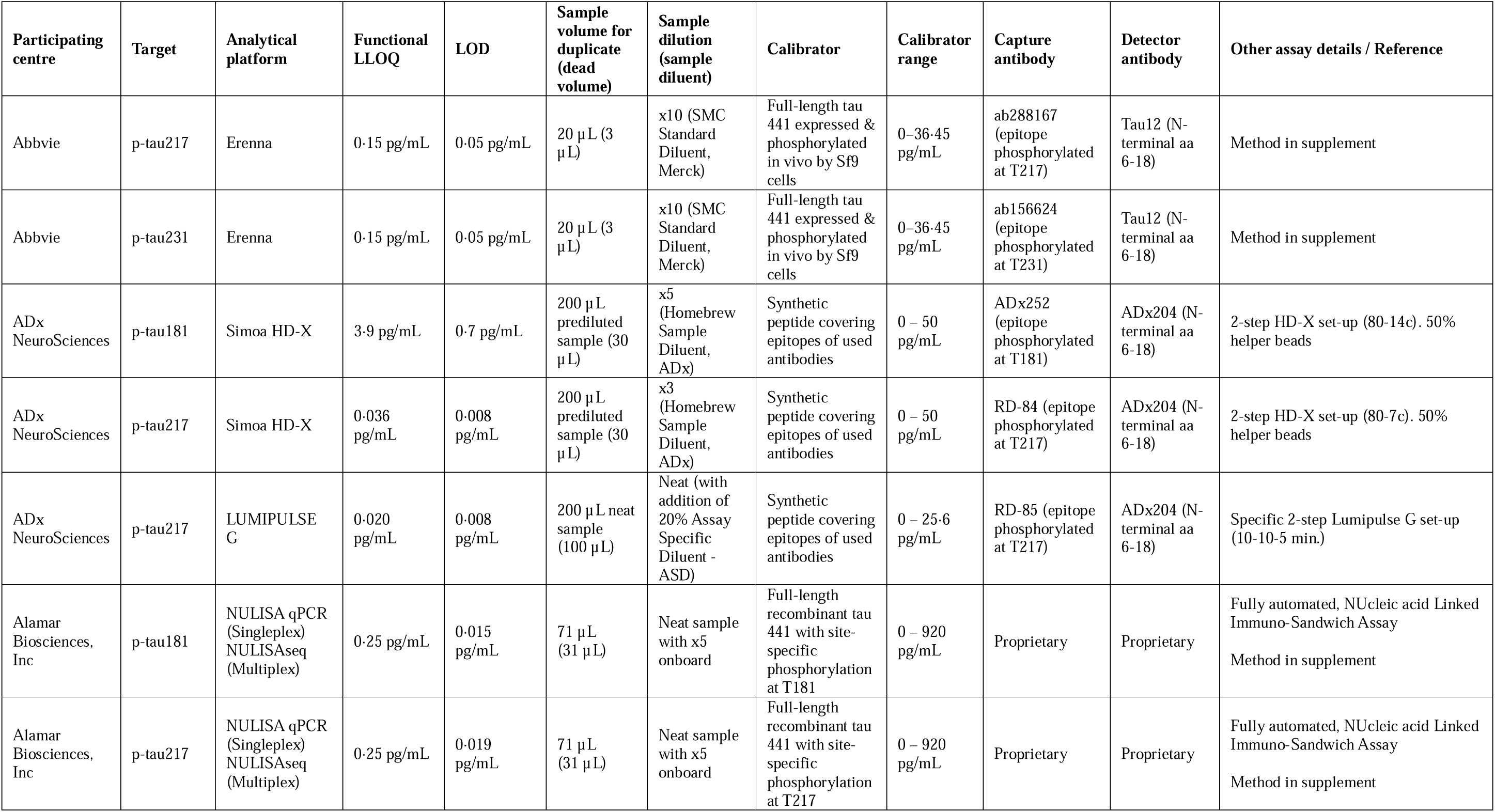
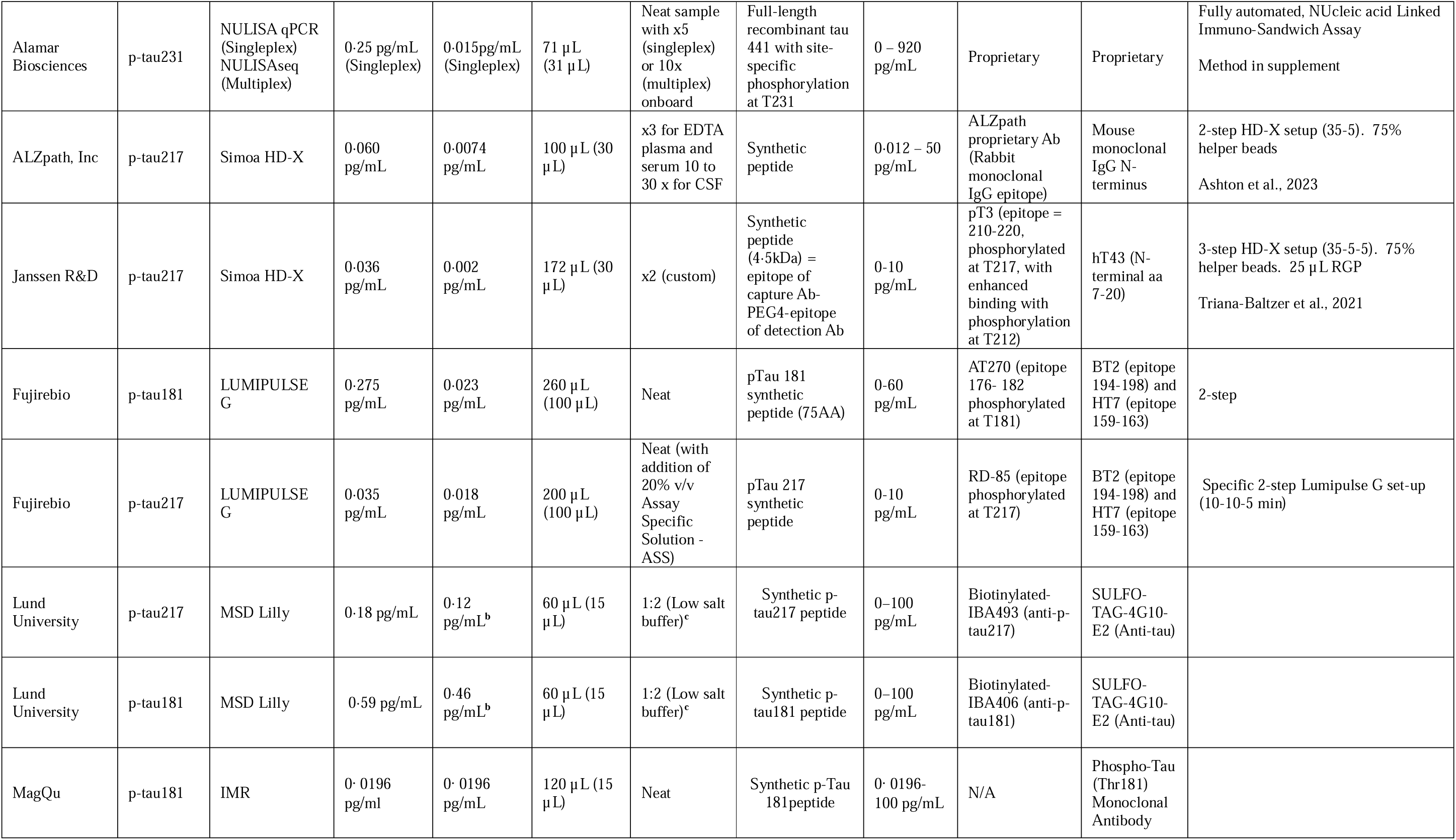
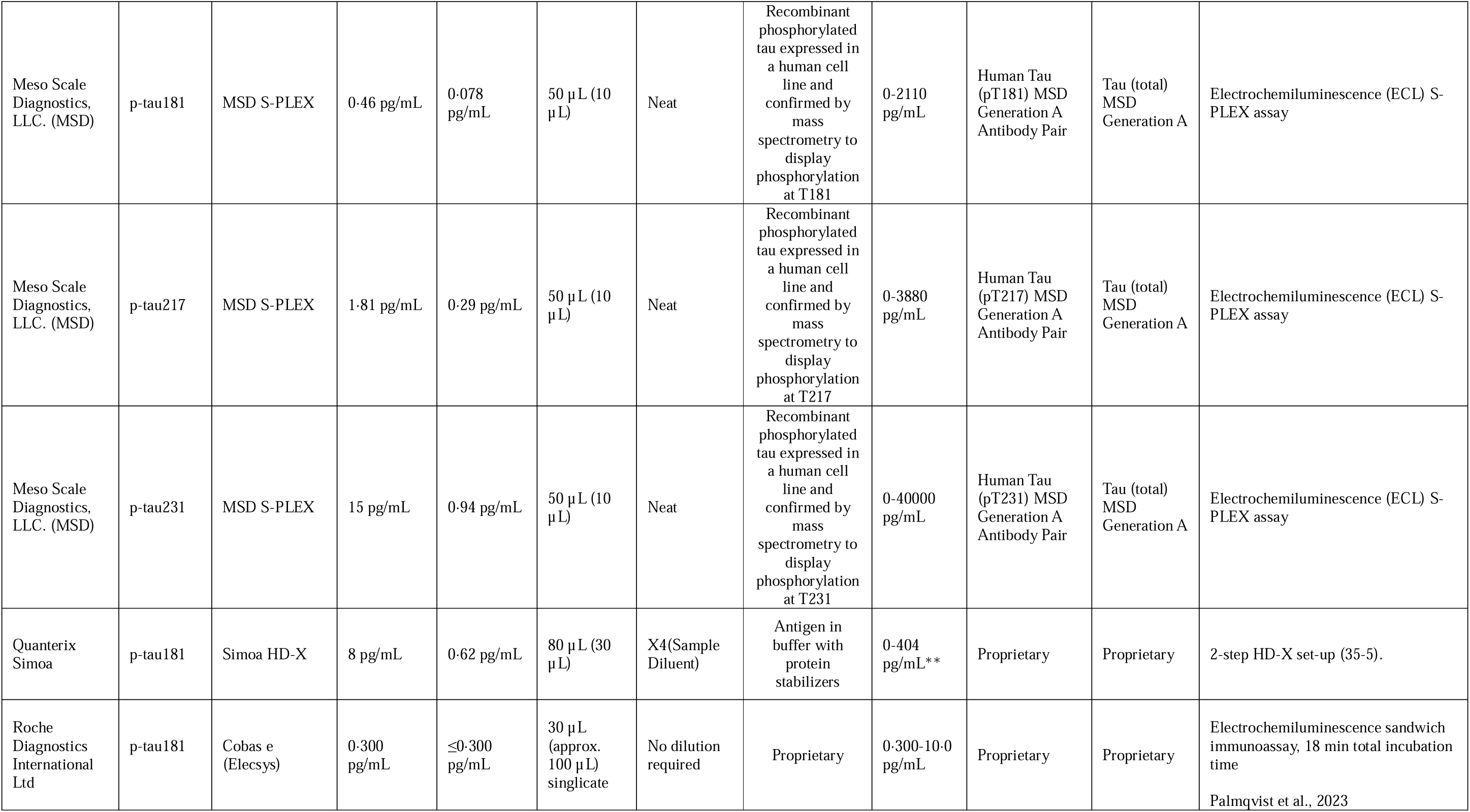
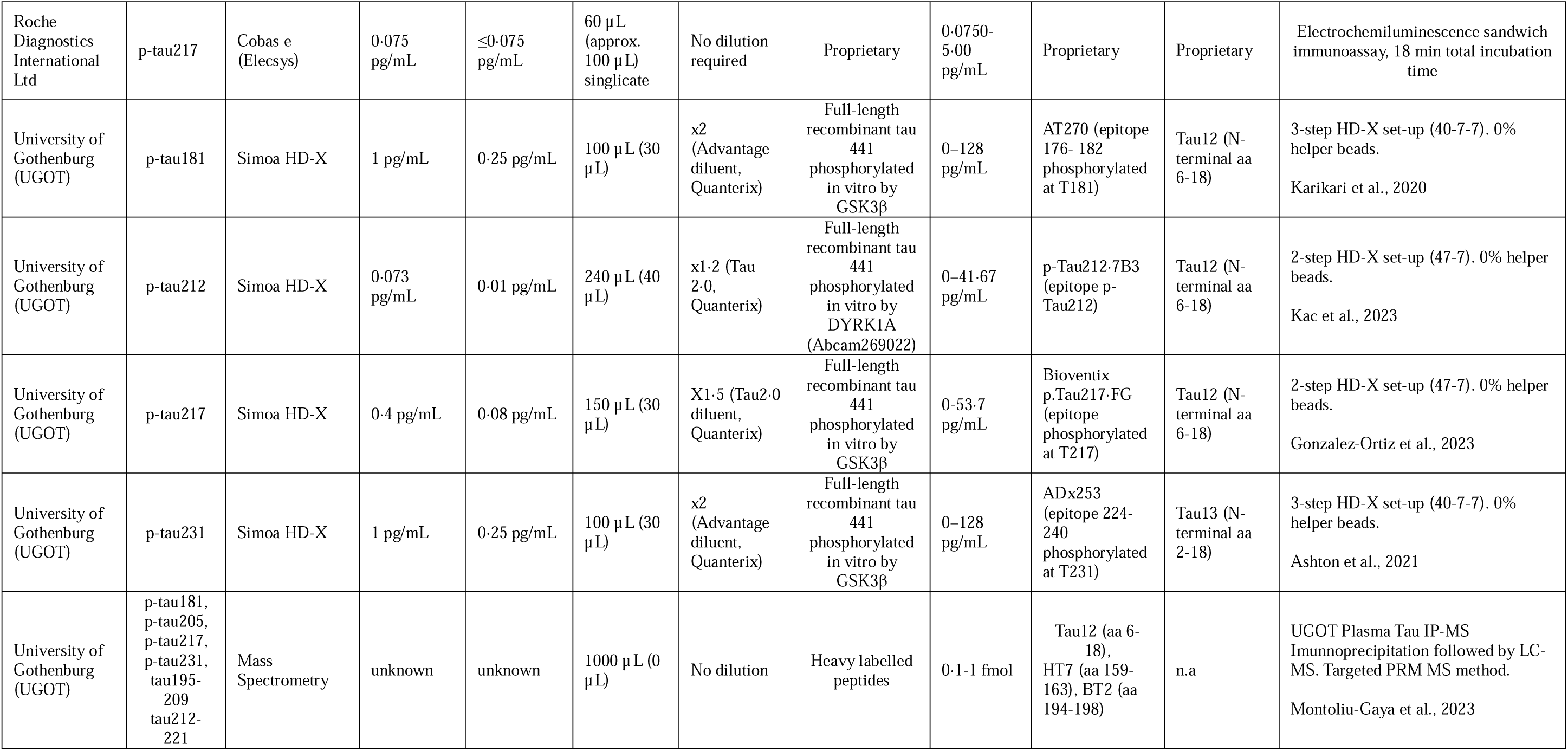
Plasma assays characteristics LLOQ=lower limit of quantification; LOD=limit of detection; n.a.=not applicable

### Candidate certified reference materials (CRM)

Each plasma assay also assessed candidate CRM created for this project. Briefly, twelve candidate CRMs (four CRMs [A-D] each at three different concentrations), were assessed (Supplementary Table 3, appendix p22). Candidate CRMs were either full-length recombinant tau1–441 phosphorylated in vitro by glycogen synthase kinase 3β (TO8–50FN; SignalChem, Vancouver, BC, Canada) in two buffers: Tau 2·0 Sample Diluent (Quanterix, #103847; A), phosphate-buffered saline + 0·05% Tween (B), or pooled EDTA plasma samples spiked with recombinant full-length tau (C) or human CSF (D). Concentrations of each candidate CRM were determined by the UGOT Simoa p-tau181 assay. Each analytical laboratory in the study was instructed to measure the candidate CRM in duplicate and to treat them as unknown plasma samples.

### Statistical analysis

Demographic information was summarised using descriptive statistics. To evaluate the magnitude of biomarker increases in the AD vs non-AD groups, mean and median fold change were computed for each plasma and CSF biomarker assays, with forest plots showing the associated standard errors. The discriminative ability of a given plasma or CSF biomarker to detect confirmed AD pathology (using an AD CSF profile as the reference standard) was evaluated by computing the area under the receiver operating characteristics curve (AUC) and visualized with forest plots alongside 95% confidence intervals. To evaluate the associations between different assays for a given p-tau biomarker (eg, correlations between different p-tau217 assays), we generated scatterplots alongside the between-assay Spearman correlation coefficient and the Passing-Bablok equation. For assays with available results in both plasma and CSF, we evaluated the cross-matrix associations with Spearman correlation, calculated both in all patients and in the AD group. A two-sided alpha of 0·05 was considered statistically significant. No multiple comparison adjustments were made, and the findings were interpreted accordingly. No CSF quantification of any participant sample fell below the limit of detection (LOD) for any assay. In plasma, measurements below the LOD were observed only for the Lilly p-tau217 assay (n=7). They were handled as previously described,^48^ by setting them to the LOD for this assay. In line with previous work with this assay, all (n=7; 100%) of the observations occurred within the non-AD CSF profile group. Where values returned as unable to be quantified, these individual sample results were omitted from the correlation analyses involving that assay alone. For candidate CRMs for plasma p-tau217, the same approach of setting observations eventually falling below the LOD to the LOD value was followed. Several p-tau217 assays presented values below the LOD for candidate CRMs as follows: ADx Lumipulse (A: n=1, B: n=2), ADx Simoa (B: n=2), Janssen Simoa (A: n=1, B: n=4), Lilly MSD (A: n=1, B: n=2), MSD S-Plex (B: n=2), and Roche Elecsys (A: n=1, B: n=2, C: n=1, D: n=1).The 95% prediction interval (PI) of the Passing-Bablok regression was calculated to conclude whether the assessed CRM were commutable with the clinical individual samples based on the positions of their values with respect to the PI. All statistical analyses were performed in R v·4·2·1 (www.r-project.org).

## RESULTS

### Participant characteristics

Out of 75 possible participant samples, 40 were selected as having enough CSF and plasma volume available, with a wide range of clinical routine CSF p-tau181 concentrations (20 to 295 pg/mL). Among these 40 participants (mean [SD] age, 63·8 [5·9] years; n [%] 17 females [42·5%]) (Table 2), 25 were categorized as having AD pathology and 15 as non-AD pathology.

**Table 2.** Participant characteristics.

|  | **All (n = 40)** | **CSF AD pathology (n = 25)** | **CSF non-AD pathology (n = 15)** |
| --- | --- | --- | --- |
| Mean age (sd), years | 63.8 (5.8) | 64.6 (6.1) | 63.8 (5.4) |
| Female, n (%) | 17 (42.5) | 12 (48.0) | 5 (33.3) |
| Mean symptom duration (sd), months | 53.2 (30.4) | 44.0 (22.4) | 68.6 (36.3) |
| Median CSF Aβ42 (IQR), pg/mL | 315 (229, 433) | 246 (224, 316) | 507 (392, 613) |
| Median CSF Aβ40, (IQR), pg/mL | 6583 (5189, 7752) | 6584 (5292, 8906) | 6581 (5107, 7403) |
| Median CSF Aβ42/40 (IQR) | 0.045 (0.037, 0.079) | 0.035 (0.038, 0.045) | 0.083 (0.078, 0.088) |
| Median p-tau181 (IQR), pg/mL | 103 (41, 137) | 124 (104, 165) | 38 (29, 42) |
| Most recent clinical diagnosis | - | Amnesic AD Dementia (n=20), mild cognitive impairment (n=1), posterior cortical atrophy (n=2), primary progressive aphasia (n=2) | Subjective cognitive decline (n=5), frontotemporal dementia not otherwise specified (n=2), semantic variant primary progressive aphasia (n=2), non-fluent variant primary progressive aphasia (n=1), meningioma and epilepsy (n=1), alcohol-related cognitive impairment (n=1), Lewy body disease (n=1), functional cognitive syndrome (n=1), autoimmune encephalitis (n=1) |

CSF biomarker values used for participant selection were obtained using the LUMIPULSE G1200 platform in clinical routine testing.

### Group-wise differences of plasma and CSF p-tau assays

All CSF assays returned results above their respective LLOQ for all participant samples. In the case of plasma assays the following assays had missing results due to values below the LLOQ (number of samples): ADx Simoa p-tau181 (1), ADx Simoa p-tau217 (5), Janssen Simoa p-tau217 (3), ALZpath Simoa p-tau217 (1), UGOT Simoa p-tau217 (1), and UGOT Simoa p-tau212 (1). Figure 1 shows the median fold-change of p-tau biomarkers in participants with AD pathology compared to those without AD pathology. For plasma (Figure 1A), the largest median fold-changes were observed for assays targeting p-tau217. UGOT IPMS (median fold-change [SE], 5.80 [2.70]), Fujirebio Lumipulse G (5·69 [3·05]), and Lilly MSD (5·49 [2·81]) all had median fold-increases > 5, while all other p-tau217 assays had median fold-increases ranging between 2·56 and 4·56 (Supplementary Table 4, appendix p23). In general, assays targeting other p-tau epitopes had median fold-changes <3 (Supplementary Table 5, appendix p24), with the exceptions being Lilly MSD p-tau181 (3·43 [1·27]), ADx Simoa p-tau181 (3·26 [1·47]) and UGOT IPMS p-tau205 (3·04 [1·34]). Plasma p-tau ratios (p-tau217/tau217 and p-tau205/tau205) did not increase the fold-changes of the p-tau assays alone. The individual boxplots for each plasma biomarker assay are shown in Supplementary Figure 1 (p-tau217), Supplementary Figure 2 (p-tau181), and Supplementary Figure 3 (p-tau231, p-tau212, and p-tau205), appendix p5-7. In CSF (Figure 1B), the highest median fold-changes were demonstrated by assays targeting p-tau217 (fold-change range 6·98 to 9·76) but also p-tau212 (8·31 [4·35]) compared to p-tau231 (median fold-change range, 4·85–5·76) and p-tau181 (fold-change range 2·43 to 4·91, excluding MagQu). In CSF, the median fold-changes were larger than in plasma for all assays (Supplementary Figure 6, appendix p10). Nevertheless, the difference between plasma and CSF was more pronounced for p-tau181, p-tau212, and p-tau231 whereas median fold-changes for plasma and CSF p-tau217 were more comparable. The Supplement displays results by mean fold-change (Supplementary Figure 4, appendix p8; Supplementary Table 7; appendix p11) and area under the curve ROC analysis (Supplementary Figure 5, appendix p9; Supplementary Table 8 and Supplementary Table 9, appendix p27-28).

**Figure 1.**
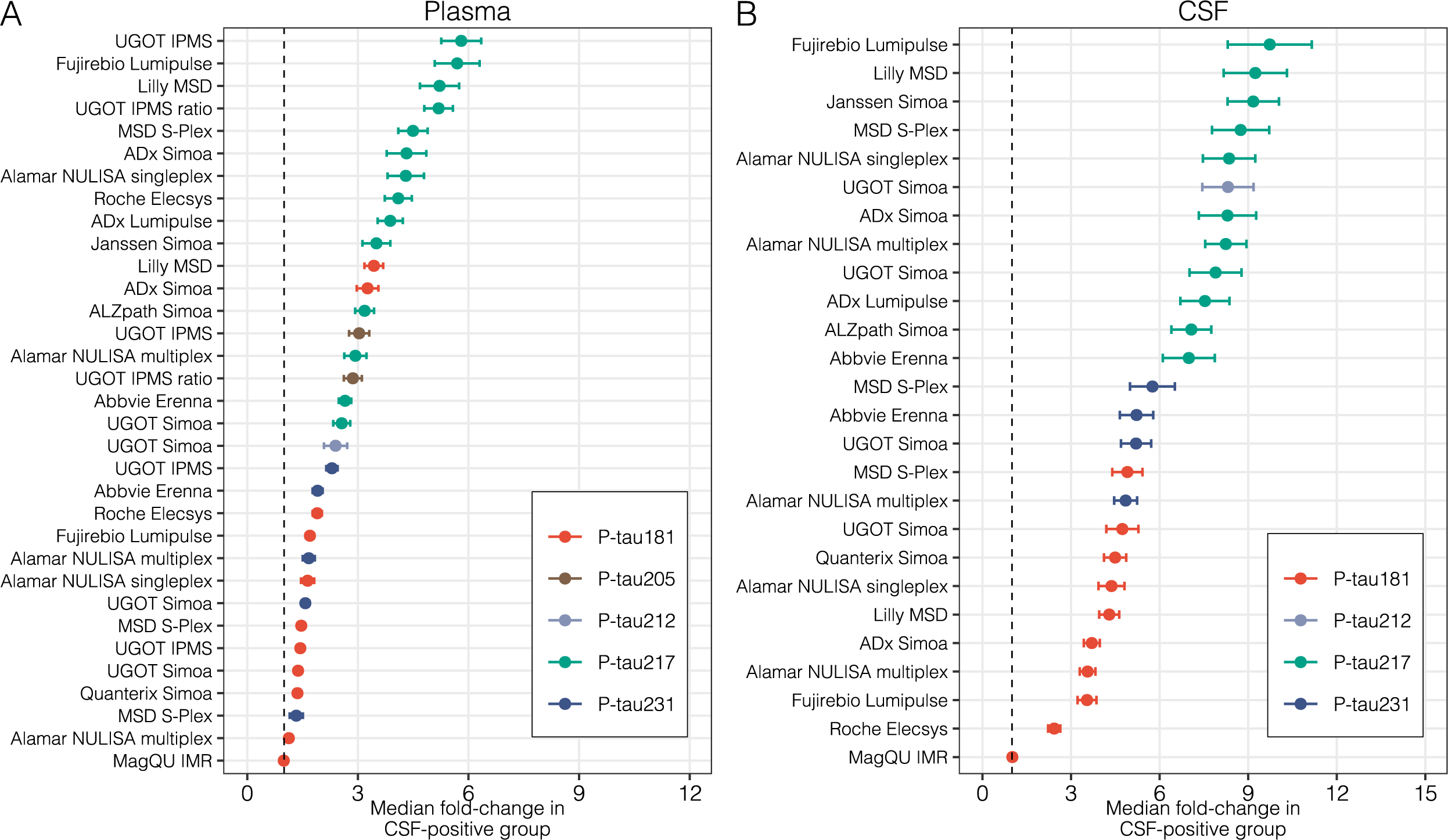
Median fold-change of plasma and CSF p-tau biomarkers in AD vs non-AD group. Forest plots indicate the median fold-change of plasma (A) and cerebrospinal fluid (CSF; B) p-tau variants in the AD pathology group compared with the non-AD pathology group. Bars correspond to standard error. Supplementary Table 4 and Supplementary Table 5 numerically describe this plot.

### Correlations between plasma assays

We examined the correlations between blood p-tau biomarker assays, grouped by phosphorylation site (Figures 2–4). A stronger overall linear relationship was observed between p-tau217 assays (mean rho=0·90; rho range 0·79 to 0·97; Figure 2), compared to p-tau181 (mean rho=0·74; rho range 0·38 to 0·91, excluding MagQu; Figure 3) and p-tau231 (mean rho=0·75; rho range 0·51 to 0·89; Figure 4).

**Figure 2.**
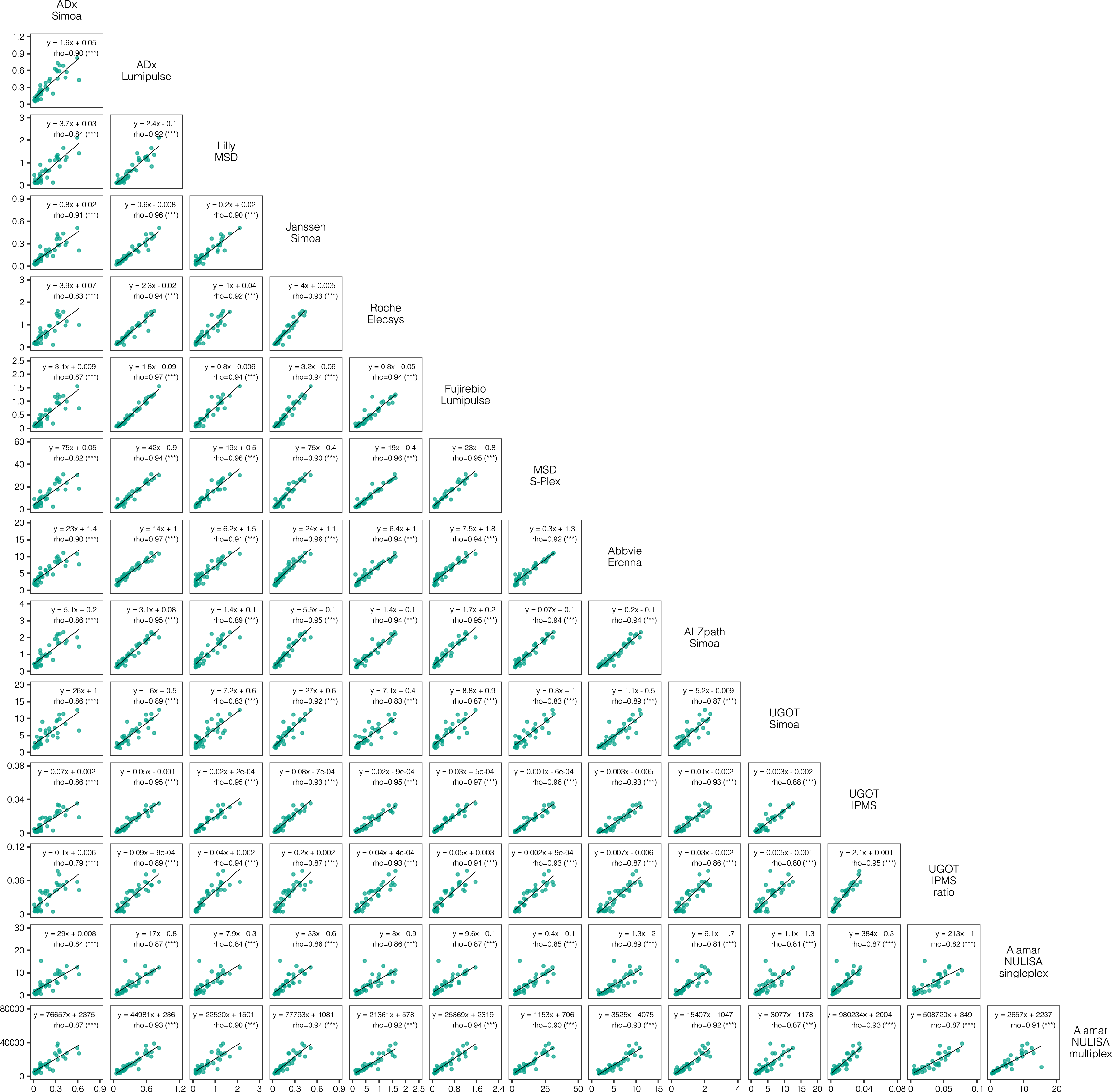
Correlations between all plasma p-tau217 assays. Scatterplots represent the continuous associations between all plasma p-tau217 assays. The dots indicate biomarker concentration and solid black line indicate the mean regression line. In each panel, text indicates the computed Passing-Bablok equation for each assay pair and the Spearman’s rho (ρ) alongside its level of statistical significance in brackets. ns= not significant, *=p<0.05, **=p<0.01, ***=p<0.0001

**Figure 3.**
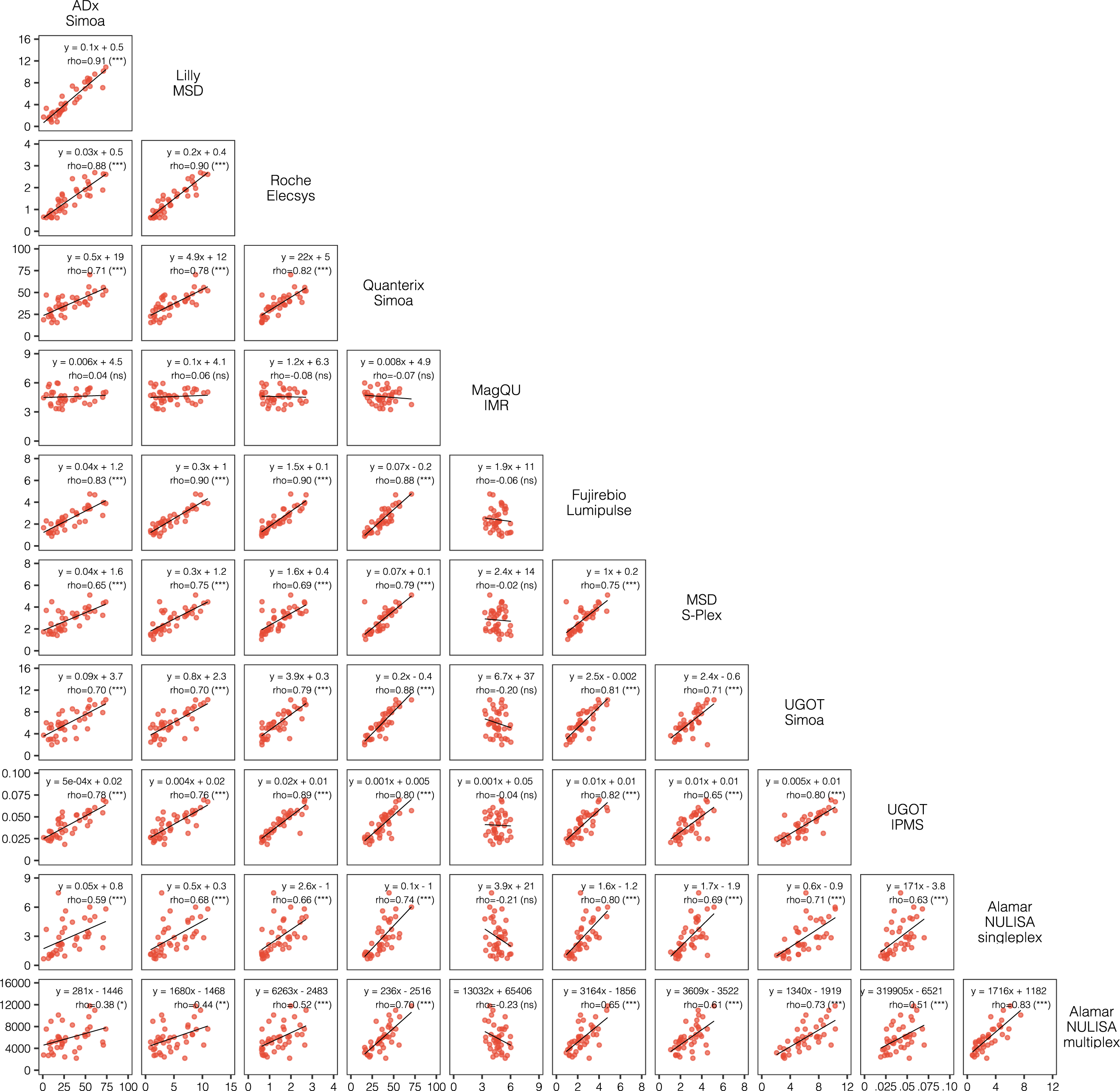
Correlations between all plasma p-tau181 assays. Scatterplots represent the continuous associations between all plasma p-tau181 assays. The dots indicate biomarker concentration and solid black line indicate the mean regression line. In each panel, text indicates the computed Passing-Bablok equation for each assay pair and the Spearman’s rho (ρ) alongside its associated level of statistical significance in brackets. ns= not significant, *=p<0.05, **=p<0.01, ***=p<0.0001

**Figure 4.**
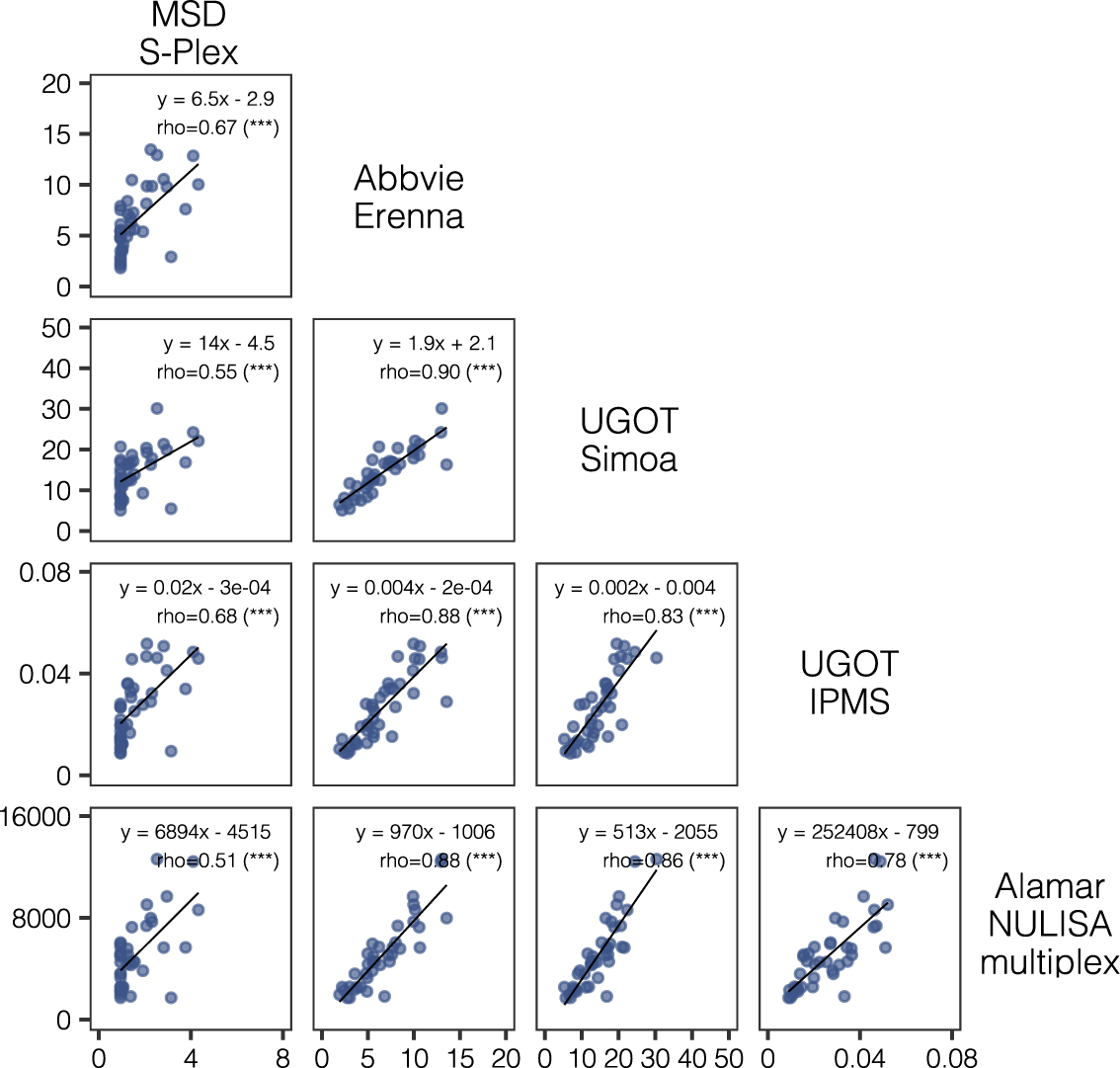
Correlations between all plasma p-tau231 assays. Scatterplots represent the continuous associations between all plasma p-tau231 assays. The dots indicate biomarker concentration and solid black line indicate the mean regression line. In each panel, text indicates the computed Passing-Bablok equation for each assay pair and the Spearman’s rho (ρ) alongside its level of statistical significance in brackets. ns= not significant, *=p<0.05, **=p<0.01, ***=p<0.0001

### Correlation between plasma and CSF

Next, we examined the correlation between plasma and CSF for the same p-tau assay. The strongest overall correspondence between plasma and CSF were observed for p-tau217 assays (Figure 5), which had a rho range of 0·61 to 0·81 (all, p*<*0·001) depending on the assay. However, when examining the AD pathology group alone, weaker, and non-significant associations where observed (rho=–0·042 to 0·36, p*>*0·05; Figure 5). The only exception was the Fujirebio Lumipulse G p-tau217 method where CSF and plasma measures were significantly correlated in the AD group (rho=0·4, p=0·048). Similar findings were observed for p-tau181 (Supplementary Figure 6, appendix p10), p-tau231 (Supplementary Figure 7, appendix p11), and p-tau212 (Supplementary Figure 8, appendix p12) with weaker and non-significant correlations in the AD pathology group. Finally, we compared our plasma biomarker assays to the CSF p-tau reference of this study (FDA-approved Lumipulse G pTau181 in CSF (Supplementary Figure 9, appendix p13). In the whole group, 31 of 33 plasma assays significantly correlated with Lumipulse G CSF p-tau181 (Supplementary Figure 9A), however, in the AD pathology group alone, no assay significantly correlated with the reference CSF p-tau181 biomarker (Supplementary Figure 9B).

**Figure 5.**
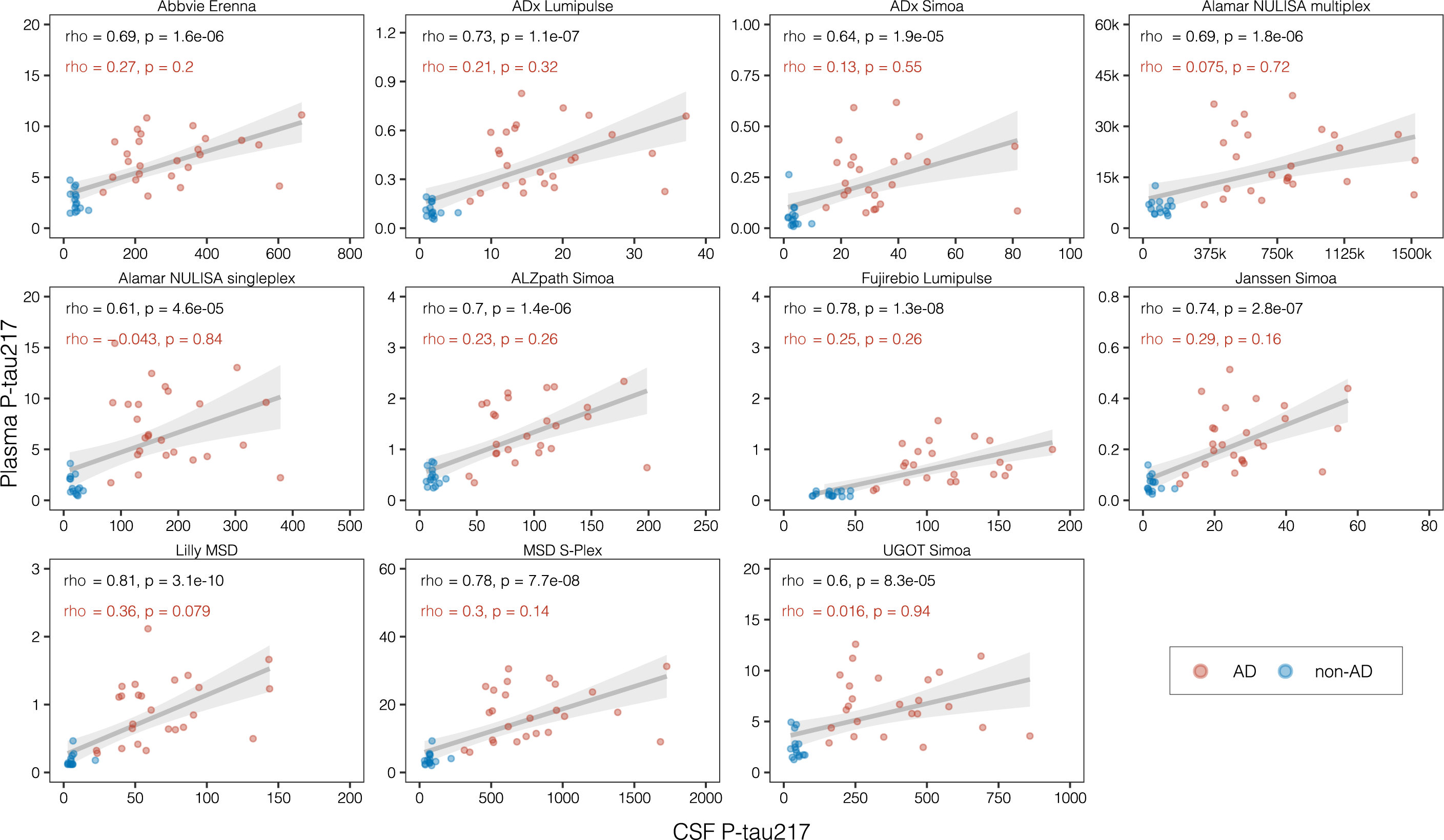
Intra-assay correlations between plasma and CSF p-tau217 biomarkers. Scatterplots represent the associations between biomarker measurements performed with the same assay in plasma (y-axis) and cerebrospinal fluid (CSF; x-axis), alongside the mean regression line with 95% confidence intervals, computed based on data from all the participants in the cohort. Red dots indicate participants from the AD group and blue dots indicate participants from the non-AD group, as defined per clinical evaluation and CSF Aβ42/Aβ40 status. In each panel, the text in black indicates the Spearman’s correlation coefficient for the entire cohort and associated p-value, with red text indicating the Spearman’s correlation coefficient and associated p-value for the AD group only.

### Candidate certified reference materials (CRM)

A total of 25 plasma assays completed the measurement of the candidate CRM. Given the clear superiority of plasma p-tau217 in the literature and this study, we report the commutability of the candidate CRMs for only those 10 p-tau217 assays that completed CRM measurement (Supplementary Figures 10 and 11, appendix p14-15). In general, all candidate CRMs were not commutable (eg, falling outside the 95% PI) for almost all method comparisons. No commutability was shown for p-tau181 or p-tau231 plasma assays (data not shown).

## DISCUSSION

The specificity of plasma p-tau to the pathologies underpinning AD^49^ offers great potential for its use as means of establishing a molecular diagnosis. Importantly, an accurate and scalable tool for determining aetiology will aid in improving clinical management by enhancing the differential diagnosis process and will reduce the need more expensive and/or invasive position emission tomography (PET) scans or lumbar punctures. In this context, it is crucial to assess the performance of different p-tau epitopes and compare different assays not only to each other but also to validated cerebrospinal fluid (CSF) markers of AD.

In this study, we performed a blinded comparison and commutability study of an unprecedented number (n=33) of different plasma p-tau measurements, including seven different p-tau epitopes or p-tau occupancy sites utilising eight antibody-based platforms. While the areas under the curve (AUCs) for all p-tau217 assays were 0·94-1, another important metric for clinical use is the median fold-change between two relevant groups (eg, distinguishing AD from non-AD pathology). Our findings clearly show that plasma p-tau217, regardless of analytical method, had larger fold-changes for determining the presence of AD pathology than p-tau181, p-tau205, p-tau212 and p-tau231. There was, however, variability in the median fold-change across p-tau217 assays. The UGOT IPMS p-tau217, which simultaneously quantifies multiple p-tau isoforms,^14^ Fujirebio Lumipulse G p-tau217, a fully-automated chemiluminescent method, and the Lilly MSD p-tau217,^38^ a manual electrochemiluminescence method each provided >5-fold median increase in the AD pathology group. This was closely followed by the MSD S-plex p-tau217, ADx Simoa p-tau217, Alamar NULISA^TM^ p-tau217, a nucleic acid linked immunoassay, and Roche Elecsys p-tau217, a fully automated electrochemiluminescence method, which showed a median >4-fold increase. The immunocapture diversity of these high-performing methods demonstrates that the analytical method is not a critical factor in determining diagnostically accurate plasma p-tau measures. In addition, for the first time, two fully automated random-access immunoassays for p-tau217 (Fujirebio Lumipulse G and Roche Elecsys), with shorter incubation times for higher analytical throughput, also produced top-tier results in this study. Again, this points towards the importance of the composition and specificity of the assay design rather than the sensitivity of the ultra-sensitive analytical platforms. This difference in fold-increase between p-tau217 tests is potentially important, given the Alzheimer’s Association guidelines for blood biomarkers ^50^ and the recent proposal for a two-step workflow for the clinical implementation of blood biomarkers.^51^ This proposal acknowledges that a binary cut-off for AD plasma biomarkers would likely obtain sub-optimal results and unacceptable numbers of false positives and false negatives.^52^ In a two-step method, which identifies high-risk and low-risk individuals based on a risk model centred around p-tau217, a biomarker with a larger fold-change will make interpretation easier and likely reduce numbers in an “intermediate” group who would need confirmatory testing with CSF or PET imaging. An assay with a larger fold-change will also be less suspectable to confounding factors^53^ impacting on its diagnostic value.

We also used the same plasma biomarker assays to measure their CSF p-tau counterparts. Previous studies have reported equivalence of diagnostic accuracy between CSF and plasma p-tau assays.^18,37^ Here, AUCs for AD pathology were also similar between plasma and CSF assays. Nevertheless, plasma p-tau showed substantially lower fold-changes for all biomarkers compared to their fold-changes in CSF, which is expected given the proximity to the diseased organ. This stresses the need for considering aspects beyond AUC values when evaluating biomarker performance and diagnostic accuracy or when choosing a biomarker for local clinical implementation. In CSF, p-tau217 and p-tau212 showed larger fold-changes compared to the other moieties. However, there was a narrower difference between plasma and CSF fold-changes for p-tau217 compared to those seen for p-tau181, p-tau212 and p-tau231, suggesting that plasma p-tau217 may more accurately reflect its CSF counterpart than other p-tau markers, and thus, brain pathology.

The strong linear correlations between blood p-tau217 assays, spanning multiple analytical platforms and assays designs, are notable and provide the potential to transition between assays, merge clinical datasets, and standardize the assays to each other using a CRM. However, in the AD pathology group alone, the associations were relatively poor, where correlations were non-significant (except for one assay) and showed, at best, a Spearman’s correlation coefficient of 0·4. Of note, it is important to bear in mind the limitations of a small sample size (n=25 in the AD group). This was also observed when correlating all plasma biomarker assay measures to the reference standard CSF assay for p-tau181 in this study. The apparent disconnect between plasma and CSF, for all assays, typically arose from higher-than-expected levels of plasma p-tau in relation to the quantified CSF measurement and suggests that alternative mechanisms (*eg*, blood-brain barrier impairment) may allow p-tau to enter the bloodstream in an advanced disease stage that is independent of the amyloid and tau brain burden. Therefore, caution must be taken not to over interpret the meaning of absolute values of plasma p-tau. Peripheral factors may also come into play in increased plasma p-tau levels, and unexpectedly high plasma p-tau values can also be observed in a single timepoint in healthy individuals followed over several weeks ^23^ and in N-terminal assay designs. Other more brain-specific tau biomarkers such as brain-derived tau (BD-tau)^12^ or assays that are more reflective of tau pathology^13,54,55^ may provide further information in this context.

This study is not free from limitations. We fully acknowledge that the sample size is not sufficient to draw clear conclusions on diagnostic superiority (particularly amongst p-tau217 assays), which was not a main aim of the study. We did not select patients based on disease severity, nor did our participants undergo tau-PET quantification, precluding us from examining whether some p-tau biomarkers or assay designs are more associated with advancing disease severity or neurofibrillary tangle pathology.^56^ Finally, the commutability aspect of the study was preliminary and designed prior to the inclusion of several different technologies in this rapidly developing field. Nonetheless, the lack of commutability of the candidate CRMs prepared here suggests that greater attention must be given to the details when developing and evaluating candidate CRMs, which will likely need to be phospho-form-specific. The strong correlations between the different p-tau217 assays suggest that further standardization work should focus on this marker and is likely to be successful.

The Alzheimer’s community can now call upon several plasma biomarker assays that can detect p-tau forms in blood which strongly indicate the presence of AD pathology. This study, of the largest number of p-tau assays to date, provides more evidence that assays targeting p-tau217 using several different methodologies show good agreement with one another, and consistently demonstrate greater fold change in AD vs non-AD groups than those targeting other p-tau forms. These results show that this is not fundamentally predicated on a single analytical platform, nor on assay design. These findings confirm that plasma p-tau217 may have clinical utility in determining the presence or absence of AD pathology in symptomatic individuals, which is relevant in the era of disease-modifying therapies.

## CONTRIBUTORS

NJA, AK, JMS and HZ conceived the study. HZ acquired funding for the study. JS provided participant samples. NJA and AK curated the data. NJA, AK and WSB analysed the data. NJA and AK drafted the initial manuscript. WSB drafted the figures. All authors reviewed and edited the manuscript.

## DATA SHARING STATEMENT

Data available: Yes

Data types: Deidentified participant data

How to access data: Requests should be directed to the corresponding authors. When available: With publication

Supporting Documents Document types: None

Who can access the data: Anonymized data will be shared by request from a qualified academic investigator.

Types of analyses: Data will be shared for the sole purpose of replicating procedures and results.

Mechanisms of data availability: Data will be available after approval of a proposal and with a signed data access agreement.

## DISCLAIMER

COBAS and ELECSYS are trademarks of Roche. All other trademarks are property of their respective owners.

The Elecsys Phospho-Tau (181P) CSF immunoassay is approved for clinical use. The Elecsys p-Tau181 and p-Tau217 prototype plasma immunoassays are not currently approved for clinical use or commercially available.

## Supporting information

STROBE statement

## Data Availability

Data available: Yes
Data types: Deidentified participant data
How to access data: Requests should be directed to the corresponding authors.
When available: With publication
Supporting Documents Document types: None
Who can access the data: Anonymized data will be shared by request from a qualified academic investigator.
Types of analyses: Data will be shared for the sole purpose of replicating procedures and results.
Mechanisms of data availability: Data will be available after approval of a proposal and with a signed data access agreement.

## ACKNOWLEGEMENT

This work was supported by the Alzheimer’s Association.

O.H. is supported by the National Institute of Aging (R01AG083740), European Research Council (ADG-101096455), Alzheimer’s Association (ZEN24-1069572, SG-23-1061717), GHR Foundation, Swedish Research Council (2022-00775), ERA PerMed (ERAPERMED2021-184), Knut and Alice Wallenberg foundation (2022-0231), Strategic Research Area MultiPark (Multidisciplinary Research in Parkinson’s disease) at Lund University, Swedish Alzheimer Foundation (AF-980907), Swedish Brain Foundation (FO2021-0293), Parkinson foundation of Sweden (1412/22), Cure Alzheimer’s fund, Rönström Family Foundation, Konung Gustaf V:s och Drottning Victorias Frimurarestiftelse, Skåne University Hospital Foundation (2020-O000028), Regionalt Forskningsstöd (2022-1259) and Swedish federal government under the ALF agreement (2022-Projekt0080).

K.B. is supported by the Swedish Research Council (grant numbers 2017-00915, and 2022-00732), the Swedish Alzheimer Foundation (grant numbers AF-930351, AF-939721, and AF-968270), the Swedish Hjärnfonden (grant numbers FO2017-0243, and ALZ2022-0006), the Swedish state under the agreement between the Swedish government and the County Councils, the ALF-agreement (grant numbers ALFGBG-715986, and ALFGBG-965240), the European Union Joint Program for Neurodegenerative Disorders (grant number JPND2019-466-236), the Alzheimer’s Association 2021 Zenith Award (grant number ZEN-21-848495), and the Alzheimer’s Association 2022-2025 Grant (grant number SG-23-1038904 QC).

H.Z. is a Wallenberg Scholar and a Distinguished Professor at the Swedish Research Council supported by grants from the Swedish Research Council (#2023-00356; #2022-01018 and #2019-02397), the European Union’s Horizon Europe research and innovation programme under grant agreement No 101053962, Swedish State Support for Clinical Research (#ALFGBG-71320), the Alzheimer Drug Discovery Foundation (ADDF), USA (#201809-2016862), the AD Strategic Fund and the Alzheimer’s Association (#ADSF-21-831376-C, #ADSF-21-831381-C, #ADSF-21-831377-C, and #ADSF-24-1284328-C), the Bluefield Project, Cure Alzheimer’s Fund, the Olav Thon Foundation, the Erling-Persson Family Foundation, Stiftelsen för Gamla Tjänarinnor, Hjärnfonden, Sweden (#FO2022-0270), the European Union’s Horizon 2020 research and innovation programme under the Marie Skłodowska-Curie grant agreement No 860197 (MIRIADE), the European Union Joint Programme – Neurodegenerative Disease Research (JPND2021-00694), the National Institute for Health and Care Research University College London Hospitals Biomedical Research Centre, and the UK Dementia Research Institute at UCL (UKDRI-1003).

## References

1. Jack CR, Jr., Bennett DA, Blennow K, et al. NIA-AA Research Framework: Toward a biological definition of Alzheimer’s disease. Alzheimers Dement 2018; 14(4): 535–62.

2. Hansson O. Biomarkers for neurodegenerative diseases. Nature Medicine 2021; 27(6): 954–63.

3. Rissman RA, Langford O, Raman R, et al. Plasma Aβ42/Aβ40 and phospho-tau217 concentration ratios increase the accuracy of amyloid PET classification in preclinical Alzheimer’s disease. Alzheimer’s & Dementia 2024; 20(2): 1214–24.

4. Schindler SE, Bollinger JG, Ovod V, et al. High-precision plasma beta-amyloid 42/40 predicts current and future brain amyloidosis. Neurology 2019; 93(17): e1647–e59.

5. Nakamura A, Kaneko N, Villemagne VL, et al. High performance plasma amyloid-beta biomarkers for Alzheimer’s disease. Nature 2018; 554(7691): 249-54.

6. Keshavan A, Pannee J, Karikari TK, et al. Population-based blood screening for preclinical Alzheimer’s disease in a British birth cohort at age 70. Brain 2021; 144(2): 434–49.

7. Mattsson N, Cullen NC, Andreasson U, Zetterberg H, Blennow K. Association Between Longitudinal Plasma Neurofilament Light and Neurodegeneration in Patients With Alzheimer Disease. JAMA Neurol 2019; 76(7): 791–9.

8. Ashton NJ, Janelidze S, Al Khleifat A, et al. A multicentre validation study of the diagnostic value of plasma neurofilament light. Nat Commun 2021; 12(1): 3400.

9. Pereira JB, Janelidze S, Smith R, et al. Plasma GFAP is an early marker of amyloid-beta but not tau pathology in Alzheimer’s disease. Brain 2021; 144(11): 3505–16.

10. Benedet AL, Mila-Aloma M, Vrillon A, et al. Differences Between Plasma and Cerebrospinal Fluid Glial Fibrillary Acidic Protein Levels Across the Alzheimer Disease Continuum. JAMA Neurol 2021; 78(12): 1471–83.

11. Mattsson N, Zetterberg H, Janelidze S, et al. Plasma tau in Alzheimer disease. Neurology 2016; 87(17): 1827–35.

12. Gonzalez-Ortiz F, Turton M, Kac PR, et al. Brain-derived tau: a novel blood-based biomarker for Alzheimer’s disease-type neurodegeneration. Brain : a journal of neurology 2023; 146(3): 1152–65.

13. Lantero-Rodriguez J, Salvadó G, Snellman A, et al. Plasma N-terminal containing tau fragments (NTA-tau): a biomarker of tau deposition in Alzheimer’s Disease. Mol Neurodegener 2024; 19(1): 19.

14. Montoliu-Gaya L, Benedet AL, Tissot C, et al. Mass spectrometric simultaneous quantification of tau species in plasma shows differential associations with amyloid and tau pathologies. Nat Aging 2023; 3(6): 661–9.

15. Ashton NJ, Moseby-Knappe M, Benedet AL, et al. Alzheimer Disease Blood Biomarkers in Patients With Out-of-Hospital Cardiac Arrest. JAMA Neurol 2023; 80(4): 388–96.

16. Ashton NJ, Puig-Pijoan A, Milà-Alomà M, et al. Plasma and CSF biomarkers in a memory clinic: Head-to-head comparison of phosphorylated tau immunoassays. Alzheimers Dement 2023; 19(5): 1913–24.

17. Janelidze S, Bali D, Ashton NJ, et al. Head-to-head comparison of 10 plasma phospho-tau assays in prodromal Alzheimer’s disease. Brain : a journal of neurology 2023; 146(4): 1592–601.

18. Barthelemy NR, Salvado G, Schindler SE, et al. Highly accurate blood test for Alzheimer’s disease is similar or superior to clinical cerebrospinal fluid tests. Nat Med 2024; 30(4): 1085–95.

19. Smirnov DS, Ashton NJ, Blennow K, et al. Plasma biomarkers for Alzheimer’s Disease in relation to neuropathology and cognitive change. Acta Neuropathol 2022; 143(4): 487–503.

20. Salvado G, Ossenkoppele R, Ashton NJ, et al. Specific associations between plasma biomarkers and postmortem amyloid plaque and tau tangle loads. EMBO Mol Med 2023; 15(5): e17123.

21. Ashton NJ, Janelidze S, Mattsson-Carlgren N, et al. Differential roles of Abeta42/40, p-tau231 and p-tau217 for Alzheimer’s trial selection and disease monitoring. Nat Med 2022; 28(12): 2555–62.

22. Mila-Aloma M, Ashton NJ, Shekari M, et al. Plasma p-tau231 and p-tau217 as state markers of amyloid-beta pathology in preclinical Alzheimer’s disease. Nat Med 2022; 28(9): 1797–801.

23. Brum WS, Ashton NJ, Simrén J, et al. Biological variation estimates of Alzheimer’s disease plasma biomarkers in healthy individuals. Alzheimers Dement 2024; 20(2): 1284–97.

24. Salvado G, Horie K, Barthelemy NR, et al. Disease staging of Alzheimer’s disease using a CSF-based biomarker model. Nat Aging 2024; 4(5): 694–708.

25. Mielke MM, Hagen CE, Xu J, et al. Plasma phospho-tau181 increases with Alzheimer’s disease clinical severity and is associated with tau- and amyloid-positron emission tomography. Alzheimers Dement 2018; 14(8): 989–97.

26. Tatebe H, Kasai T, Ohmichi T, et al. Quantification of plasma phosphorylated tau to use as a biomarker for brain Alzheimer pathology: pilot case-control studies including patients with Alzheimer’s disease and down syndrome. Mol Neurodegener 2017; 12(1): 63.

27. Karikari TK, Pascoal TA, Ashton NJ, et al. Blood phosphorylated tau 181 as a biomarker for Alzheimer’s disease: a diagnostic performance and prediction modelling study using data from four prospective cohorts. Lancet Neurol 2020; 19(5): 422–33.

28. Triana-Baltzer G, Moughadam S, Slemmon R, et al. Development and validation of a high-sensitivity assay for measuring p217+tau in plasma. Alzheimers Dement (Amst) 2021; 13(1): e12204.

29. Yang CC, Chiu MJ, Chen TF, Chang HL, Liu BH, Yang SY. Assay of Plasma Phosphorylated Tau Protein (Threonine 181) and Total Tau Protein in Early-Stage Alzheimer’s Disease. J Alzheimers Dis 2018; 61(4): 1323–32.

30. Janelidze S, Mattsson N, Palmqvist S, et al. Plasma P-tau181 in Alzheimer’s disease: relationship to other biomarkers, differential diagnosis, neuropathology and longitudinal progression to Alzheimer’s dementia. Nat Med 2020; 26(3): 379–86.

31. Palmqvist S, Stomrud E, Cullen N, et al. An accurate fully automated panel of plasma biomarkers for Alzheimer’s disease. Alzheimers Dement 2023; 19(4): 1204–15.

32. Barthelemy NR, Horie K, Sato C, Bateman RJ. Blood plasma phosphorylated-tau isoforms track CNS change in Alzheimer’s disease. J Exp Med 2020; 217(11).

33. Bayoumy S, Verberk IMW, den Dulk B, et al. Clinical and analytical comparison of six Simoa assays for plasma P-tau isoforms P-tau181, P-tau217, and P-tau231. Alzheimers Res Ther 2021; 13(1): 198.

34. Schindler SE, Petersen KK, Saef B, et al. Head-to-head comparison of leading blood tests for Alzheimer’s disease pathology. medRxiv 2024.

35. Mielke MM, Frank RD, Dage JL, et al. Comparison of Plasma Phosphorylated Tau Species With Amyloid and Tau Positron Emission Tomography, Neurodegeneration, Vascular Pathology, and Cognitive Outcomes. JAMA Neurol 2021; 78(9): 1108–17.

36. Ashton NJ, Brum WS, Di Molfetta G, et al. Diagnostic Accuracy of a Plasma Phosphorylated Tau 217 Immunoassay for Alzheimer Disease Pathology. JAMA Neurol 2024.

37. Palmqvist S, Janelidze S, Quiroz YT, et al. Discriminative Accuracy of Plasma Phospho-tau217 for Alzheimer Disease vs Other Neurodegenerative Disorders. Jama 2020.

38. Palmqvist S, Janelidze S, Quiroz YT, et al. Discriminative Accuracy of Plasma Phospho-tau217 for Alzheimer Disease vs Other Neurodegenerative Disorders. JAMA 2020; 324(8): 772–81.

39. Janelidze S, Stomrud E, Smith R, et al. Cerebrospinal fluid p-tau217 performs better than p-tau181 as a biomarker of Alzheimer’s disease. Nat Commun 2020; 11(1): 1683.

40. Kivisäkk P, Carlyle BC, Sweeney T, et al. Plasma biomarkers for diagnosis of Alzheimer’s disease and prediction of cognitive decline in individuals with mild cognitive impairment. Front Neurol 2023; 14: 1069411.

41. Kivisakk P, Fatima HA, Cahoon DS, et al. Clinical evaluation of a novel plasma pTau217 electrochemiluminescence immunoassay in Alzheimer’s disease. Sci Rep 2024; 14(1): 629.

42. Bayoumy S, Verberk IMW, den Dulk B, et al. Clinical and analytical comparison of six Simoa assays for plasma P-tau isoforms P-tau181, P-tau217, and P-tau231. Alzheimer’s Research & Therapy 2021; 13(1): 198.

43. Karikari TK, Emersic A, Vrillon A, et al. Head-to-head comparison of clinical performance of CSF phospho-tau T181 and T217 biomarkers for Alzheimer’s disease diagnosis. Alzheimers Dement 2021; 17(5): 755–67.

44. Kac PR, Gonzalez-Ortiz F, Emersic A, et al. Plasma p-tau212: antemortem diagnostic performance and prediction of autopsy verification of Alzheimer’s disease neuropathology. medRxiv 2023.

45. Gonzalez-Ortiz F, Ferreira PCL, Gonzalez-Escalante A, et al. A novel ultrasensitive assay for plasma p-tau217: Performance in individuals with subjective cognitive decline and early Alzheimer’s disease. Alzheimers Dement 2023.

46. Ashton NJ, Pascoal TA, Karikari TK, et al. Plasma p-tau231: a new biomarker for incipient Alzheimer’s disease pathology. Acta Neuropathol 2021; 141(5): 709–24.

47. Ashton NJ, Benedet AL, Pascoal TA, et al. Cerebrospinal fluid p-tau231 as an early indicator of emerging pathology in Alzheimer’s disease. EBioMedicine 2022; 76: 103836.

48. Janelidze S, Palmqvist S, Leuzy A, et al. Detecting amyloid positivity in early Alzheimer’s disease using combinations of plasma Aβ42/Aβ40 and p-tau. Alzheimers Dement 2022; 18(2): 283–93.

49. Lantero Rodriguez J, Karikari TK, Suarez-Calvet M, et al. Plasma p-tau181 accurately predicts Alzheimer’s disease pathology at least 8 years prior to post-mortem and improves the clinical characterisation of cognitive decline. Acta Neuropathol 2020; 140(3): 267–78.

50. Hansson O, Edelmayer RM, Boxer AL, et al. The Alzheimer’s Association appropriate use recommendations for blood biomarkers in Alzheimer’s disease. Alzheimers Dement 2022; 18(12): 2669–86.

51. Brum WS, Cullen NC, Janelidze S, et al. A two-step workflow based on plasma p-tau217 to screen for amyloid beta positivity with further confirmatory testing only in uncertain cases. Nat Aging 2023; 3(9): 1079–90.

52. Figdore DJ, Griswold M, Bornhorst JA, et al. Optimizing cutpoints for clinical interpretation of brain amyloid status using plasma p-tau217 immunoassays. Alzheimers Dement 2024.

53. Mielke MM, Dage JL, Frank RD, et al. Performance of plasma phosphorylated tau 181 and 217 in the community. Nat Med 2022; 28(7): 1398–405.

54. Lantero-Rodriguez J, Tissot C, Snellman A, et al. Plasma and CSF concentrations of N-terminal tau fragments associate with in vivo neurofibrillary tangle burden. Alzheimers Dement 2023; 19(12): 5343–54.

55. Horie K, Salvado G, Barthelemy NR, et al. CSF MTBR-tau243 is a specific biomarker of tau tangle pathology in Alzheimer’s disease. Nat Med 2023; 29(8): 1954–63.

56. Mattsson-Carlgren N, Collij LE, Stomrud E, et al. Plasma Biomarker Strategy for Selecting Patients With Alzheimer Disease for Antiamyloid Immunotherapies. JAMA Neurol 2024; 81(1): 69–78.

