## Supplementary appendix for "The Alzheimer’s Association Global Biomarker Standardization Consortium (GBSC) plasma phospho-tau Round Robin study"

^1^ Department of Psychiatry and Neurochemistry, Institute of Neuroscience & Physiology, the Sahlgrenska Academy at the University of Gothenburg, Mölndal, Sweden; ^2^ King's College London, Institute of Psychiatry, Psychology and Neuroscience Maurice Wohl Institute Clinical Neuroscience Institute London, UK; ^3^ NIHR Biomedical Research Centre for Mental Health and Biomedical Research Unit for Dementia at South London and Maudsley NHS Foundation London, UK; ^4^ Centre for Age-Related Medicine, Stavanger University Hospital, Stavanger, Norway; ^5^ Dementia Research Centre, UCL Queen Square Institute of Neurology, University College London, London, UK.; ^6^ Graduate Program in Biological Sciences: Biochemistry, Universidade Federal do Rio Grande do Sul (UFRGS), Porto Alegre, Brazil; ^7^ AbbVie Deutschland GmbH & Co. KG, Neuroscience Research, Knollstrasse, 67061 Ludwigshafen, Germany; ^8^ ADx NeuroSciences N.V., Technologiepark 6, 9052 Ghent, Belgium; ^9^ Alamar Biosciences, Inc., Fremont, CA, USA; ^10^ ALZpath Inc., Carlsbad, CA, USA; ^11^ Fujirebio Europe N.V., Ghent, Belgium; ^12^ Enigma Biomedical Group, CA, USA ^13^ Neuroscience Biomarkers, Janssen Research and Development, La Jolla, California, USA; ^14^ Clinical Memory Research Unit, Department of Clinical Sciences, Lund University, Lund 22184, Sweden; ^15^ MagQu Co., Ltd., New Taipei City 112, Taiwan; ^16^ Meso Scale Diagnostics, LLC., Rockville, Maryland, USA; ^17^ Quanterix Corp, Billerica, MA, USA; ^18^ Roche Diagnostics GmbH, Penzberg, Germany; ^19^ Clinical Neurochemistry Laboratory, Sahlgrenska University Hospital, Mölndal, Sweden; ^20^ Memory Clinic, Skåne University Hospital, Malmö 20502, Sweden; ^21^ Alzheimer's Therapeutic Research Institute, Keck School of Medicine of the University of Southern California, San Diego, CA 92121, USA; ^22^ Division of Medical & Scientific Relations, Alzheimer's Association, Chicago, Illinois, USA; ^23^ Department of pathology & laboratory Medicine, Perelman School of Medicine, University of Pennsylvania, Philadelphia, PA 19104; ^24^ Paris Brain Institute, ICM, Pitié-Salpêtrière Hospital, Sorbonne University, Paris, France; ^25^ Neurodegenerative Disorder Research Center, Division of Life Sciences and Medicine, and Department of Neurology, Institute on Aging and Brain Disorders, University of Science and Technology of China and First Affiliated Hospital of USTC, Hefei, P.R. China; ^26^ UK Dementia Research Institute, University College London, London, UK; ^27^ Department of Neurodegenerative Disease, UCL Institute of Neurology, Queen Square, London, UK; ^28^ Hong Kong Center for Neurodegenerative Diseases, Science Park, Hong Kong, China; ^29^ School of Medicine and Public Health, University of Wisconsin-Madison, Madison, Wisconsin, USA

^*^ Nicholas J. Ashton, Ashvini Keshavan, and Wagner S. Brum contributed equally as first authors.

^#^ Henrik Zetterberg and Jonathan M. Schott contributed equally as senior authors.

**Supplementary Methods** ………………………………………………………………………………………3

**Supplementary Methods**

**Details of unpublished methods provided by vendors**

***Abbvie p-tau217 and p-tau231***

The respective monoclonal antibodies were either biotinylated and coated to magnetic beads using the SMC Capture Reagent Labeling Kit or conjugated to Alexa647 dye using the SMC Detection Reagent Labeling Kit (both Merck) as per manufacturer’s instructions. Immunoassay analysis was carried out using the Immunoassay Development kit (Merck). In short, samples were diluted in Standard Diluent (dilution factors 50 for CSF or 10 for Plasma) prior to incubation for 2 h with the Capture antibody beads and for 1h with the Detection antibody at 25°C. All washing steps in between incubations were done on an automated plate washer (Tecan) using Wash Buffer with Proclin. Prior to elution, beads were transferred to a new 96-well plate using the Viaflo96 pipetting robot (Integra) to minimize background signals. Detection antibodies were eluted from the beads with Elution Buffer B for 20 min at 25°C. To increase robustness, the eluate was neutralized and transferred to a 384-well scanning plate with a single tip using the Viaflo96 pipetting robot. Detection of signals for thius study was performed on the Erenna (Singulex) platform but also could be performed with slight adaptations on the next generation device SMCxPro (Merck).

***ADx Lumipulse p-tau217***

N-terminal p-tau217 concentrations were measured at ADx NeuroSciences N.V. (Ghent, Belgium) using a prototype Lumipulse G pTau 217 Plasma assay on a LUMIPULSE G1200 instrument. The assay has a specific 2-step set-up where the analyte is first captured on RD-85 (p-tau217 specific mouse mAb) coated particles, and after washing is detected with alkaline phosphatase (ALP) labelled RD-73 conjugate, a CHO recombinantly expressed version of the hybridoma based N-terminal antibody ADx204. The assay includes a synthetic peptide containing the two antibody epitopes as the calibrator. Samples were tested as singlets due to high analytical repeatability of the assay. Before analysis, EDTA plasma was centrifuged for 5 mins at 2000 g. In the automated assay protocol, 100 µL neat EDTA plasma was pretreated with 25 µL assay specific diluent (20% ASD v/v) before addition to the 150 µL particle solution in the immunocartridge. After 10 mins incubation, the complex was washed and allowed to incubate for 10 mins with enzyme labeled detector mAb. After final wash, AMPPD substrate was added for 5 min to generate an analyte specific signal. For CSF analysis, the sample was prediluted (dilution factor 1·45) in particle diluent and subsequently treated as plasma sample in the automated protocol.

***ADx Simoa p-tau217***

Concentrations were measured at ADx NeuroSciences N.V. (Ghent, Belgium) using a 2-step Homebrew Simoa immuno-assay developed by ADx NeuroSciences. Samples were analyzed in duplicate using a biotinylated N-terminal antibody, ADx204, as detector and RD-84, a p-tau217-specific mouse mAb, as capture on the paramagnetic beads. The assay was calibrated with a synthetic peptide representing the antibody epitopes. Before analysis, EDTA plasma is centrifuged for 8 mins at 10,000 g and prediluted offline to 33% (dilution factor 3) in homebrew sample diluent. CSF is prediluted to 5% (dilution factor 20) in sample diluent and subsequently analyzed as the plasma samples; 100 µL of prediluted sample is incubated for 60 mins (80 candences) with 25 µL capture beads and 20 µL detector. After washing, SβG is added to the immunocomplex (5 mins, 7 cadences) for fluorescent signal generation after RGP substrate addition.

***Alamar Biosciences p-tau181, p-tau217, p-tau231***

NULISAseq assays were performed at Alamar Biosciences as described previously (Feng et al.). Briefly, plasma and CSF samples stored at -80°C were thawed on ice and centrifuged at 10,000g for 10 mins. Single-plex p-tau181 and p-tau217 NULISA qPCR assays utilized 20 µL of sample per reaction with duplicate measurements. For CNS disease panel NULISAseq, 10 µL of each sample was measured in a singlet format. A Hamilton-based automation instrument was used to perform the NULISA workflow, starting with immunocomplex formation with DNA-barcoded capture and detection antibodies. The subsequent steps involved capturing and washing immunocomplexes on paramagnetic oligo-dT beads, releasing them into a low-salt buffer, and capturing and washing on streptavidin beads. Proximal ends of DNA strands on each immunocomplex were ligated with T4 DNA ligase, generating a DNA reporter. For CNS disease panel NULISAseq assay, sample-specific barcodes were also incorporated into the DNA reporter during this step. In singleplex qPCR assays, molecule quantification was achieved through qPCR reactions. For NULISAseq, DNA reporters containing both target-specific and sample-specific barcodes were pooled and subjected to PCR amplification, followed by purification and sequencing on Illumina NextSeq 2000.

***Fujirebio Lumipulse G p-tau217***

pTau 217 levels in plasma and CSF were measured at Fujirebio Europe N.V. (Gent, Belgium) using a prototype of the Lumipulse G pTau 217 Plasma RUO assay on a LUMIPULSE G instrument. The assay works according to a specific 2-step set-up where the analyte is first captured in presence of an assay specific solution on RD85 coated particles, and after washing is detected with ALP labelled HT7 / BT2 conjugate. The assay uses a synthetic peptide containing the three epitopes as the calibrator. Samples were tested in singlicate. The assay uses a sample volume of 40 µL or 100 µL for plasma and CSF, respectively. Additionally, for CSF, the calibrator range was extended to accommodate the broader concentration range.

***Meso Scale S-PLEX p-tau231***

Ultrasensitive MSD S-PLEX assays were performed at Meso Scale Diagnostics, LLC. (Rockville, MD, USA). Samples were analyzed in duplicate on an MSD Sector S 600MM reader employing a sandwhich immunoassay format with monoclonal antibodies and electrochemiluminescence (ECL) detection. In addition to the CRMs, three MSD quality control (QC) samples spanning the assay range were run in duplicate on each plate. Calibrators for the three assays were prepared from recombinant phosphorylated tau expressed in a mammalian system and confirmed by mass spectrometry to display phosphorylation at T181, T217, and T231, respectively. A 7-point calibration curve with a blank Cal-8 was prepared by 4-fold serial dilution and run in duplicate on each plate. Plasma samples were measured in duplicate using 25μL of neat sample per well. CSF samples were measured in duplicate using 25μL of 2-fold diluted sample per well.

***Roche Elecsys*^®^ *p-tau217 (Roche Diagnostics International Ltd, Rotkreuz, Switzerland)***

Plasma samples were analyzed in singlicates using an in-house Elecsys plasma prototype immunoassay (not commercially available) for p-tau217 on a Cobas^®^ e 801 analyzer at Roche Diagnostics GmbH, Penzberg, Germany. The Elecsys method is an antibody-based technique which gives high analytical sensitivity (a sandwich immunoassay), based on one capture and one detection antibody (which increases the specificity). For that reason, it is performed on neat plasma, without any cleanup or pre-treatment step. The general Elecsys assay setup was as follows: 60 µL of sample, a biotinylated monoclonal antibody and a monoclonal antibody labeled with a ruthenium complex were first co-incubated for 9 minutes to form a sandwich complex comprising the biotinylated antibody, analyte and the ruthenylated antibody. In the second incubation step (9 minutes), streptavidin-coated microparticles (Elecsys beads) were added to the mixture of the first incubation step and, as a result, the complex comprising the biotinylated antibody, analyte and the ruthenylated antibody became bound to the solid phase via interaction of biotin and streptavidin. The reaction mixture was aspirated into the measuring cell where the microparticles were magnetically captured onto the surface of the electrode. Unbound substances were then removed with ProCell M. Application of a voltage to the electrode then induced chemiluminescent emission, which was measured by a photomultiplier. Sample concentrations were finally determined from an instrument-specific calibration curve.

**Supplementary Figure 1.** Boxplots of plasma p-tau217 levels in AD and non-AD groups.

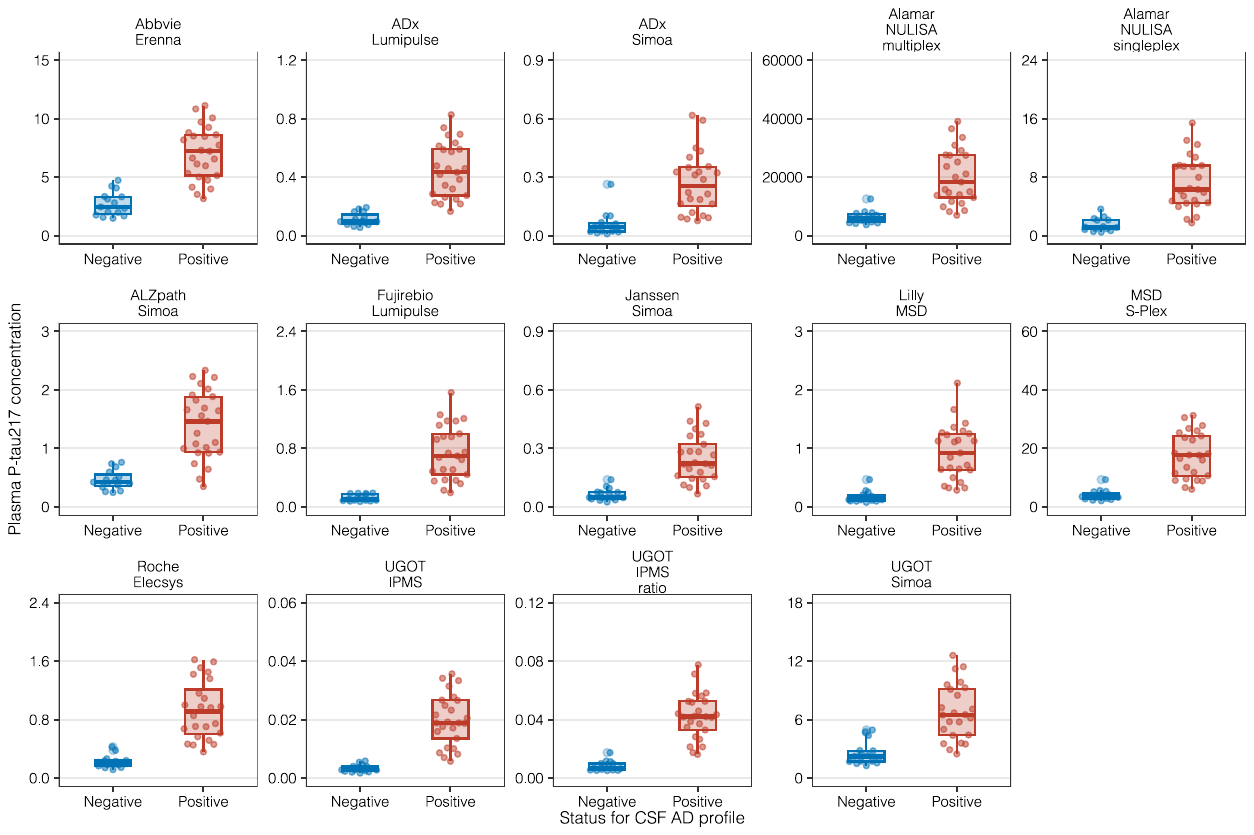

Boxplots showing the concentrations of plasma p-tau217 in pg/mL on the y-axis and CSF AD pathology status on the x-axis, for the AD pathology (red) and non-AD pathology (blue) groups. Each panel represents a different assay for plasma p-tau217. For immunoprecipitation mass-spectrometry (IPMS), p-tau217 is also reported in a ratio between p-tau217 and a non-phosphorylated peptide at tau212-221.

**Supplementary Figure 2.** Boxplots of plasma p-tau181 levels in AD and non-AD groups.

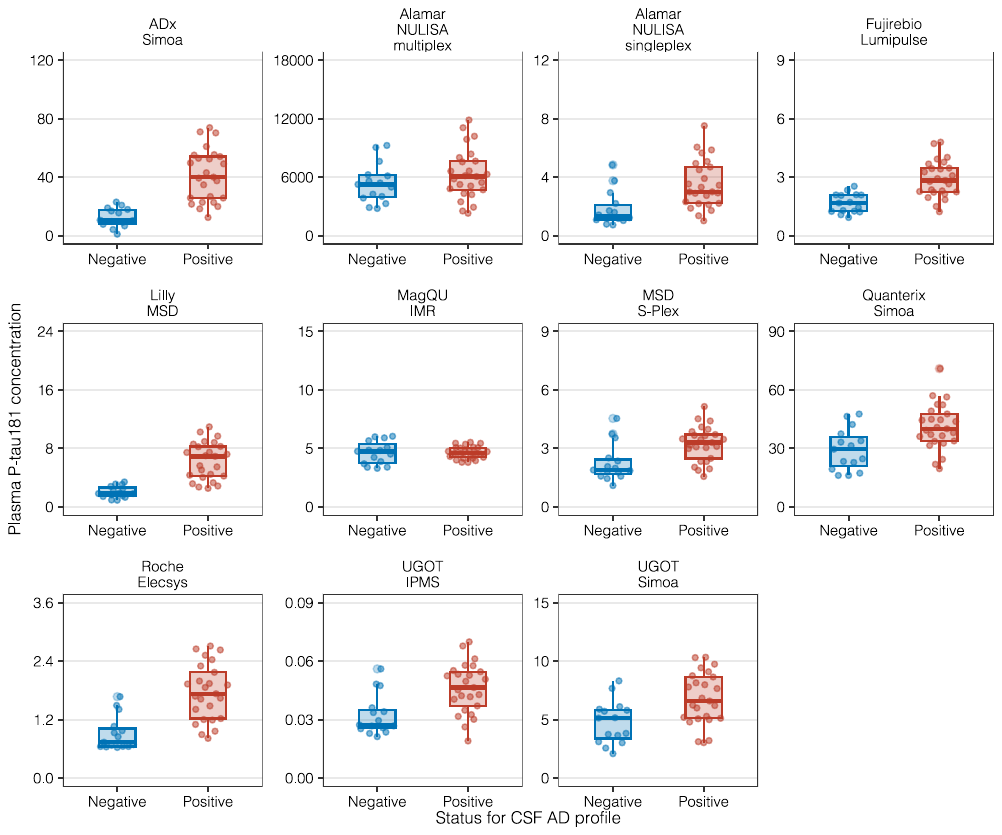

Boxplots showing the concentrations of plasma p-tau181 in pg/mL on the y-axis and CSF AD pathology status on the x-axis, for the AD pathology (red) and non-AD pathology (blue) groups. Each panel represents a different assay for plasma p-tau181.

**Supplementary Figure 3.** Boxplots of plasma p-tau231, p-tau205 and p-tau212 levels in AD and non-AD groups.

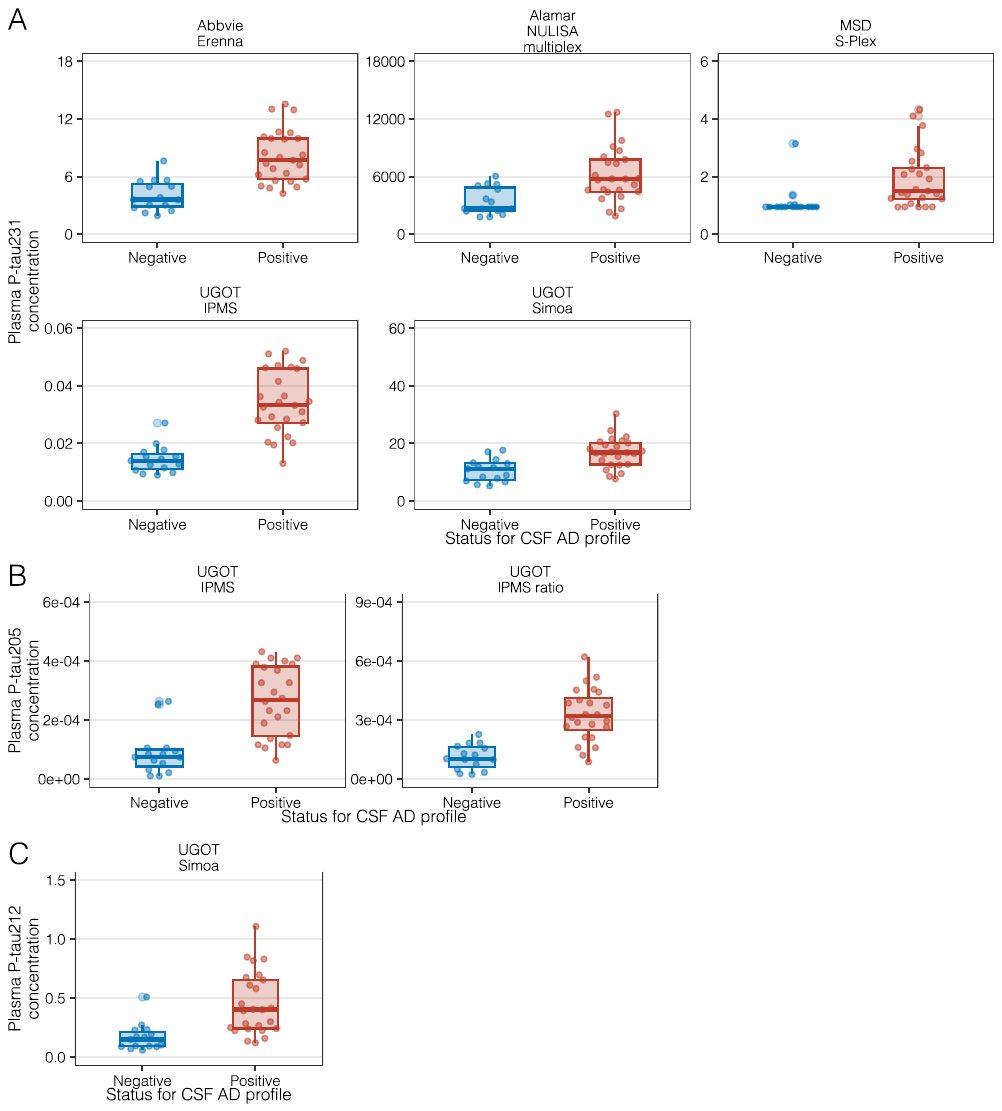

Boxplots showing the concentrations of plasma p-tau231 (A; top row), p-tau205 (B; middle row) and p-tau212 (C; bottom row) in pg/mL on the y-axis and CSF AD pathology status on the x-axis, for the AD pathology (red) and non-AD pathology (blue) groups. Each panel represents a different assay for each plasma p-tau biomarker. For immunoprecipitation mass-spectrometry (IPMS), p-tau205 is also reported in a ratio between p-tau205 and a non-phosphorylated peptide at tau195-209.

**Supplementary Figure 4.** Mean fold-change of plasma and CSF p-tau biomarkers in AD vs non-AD group.

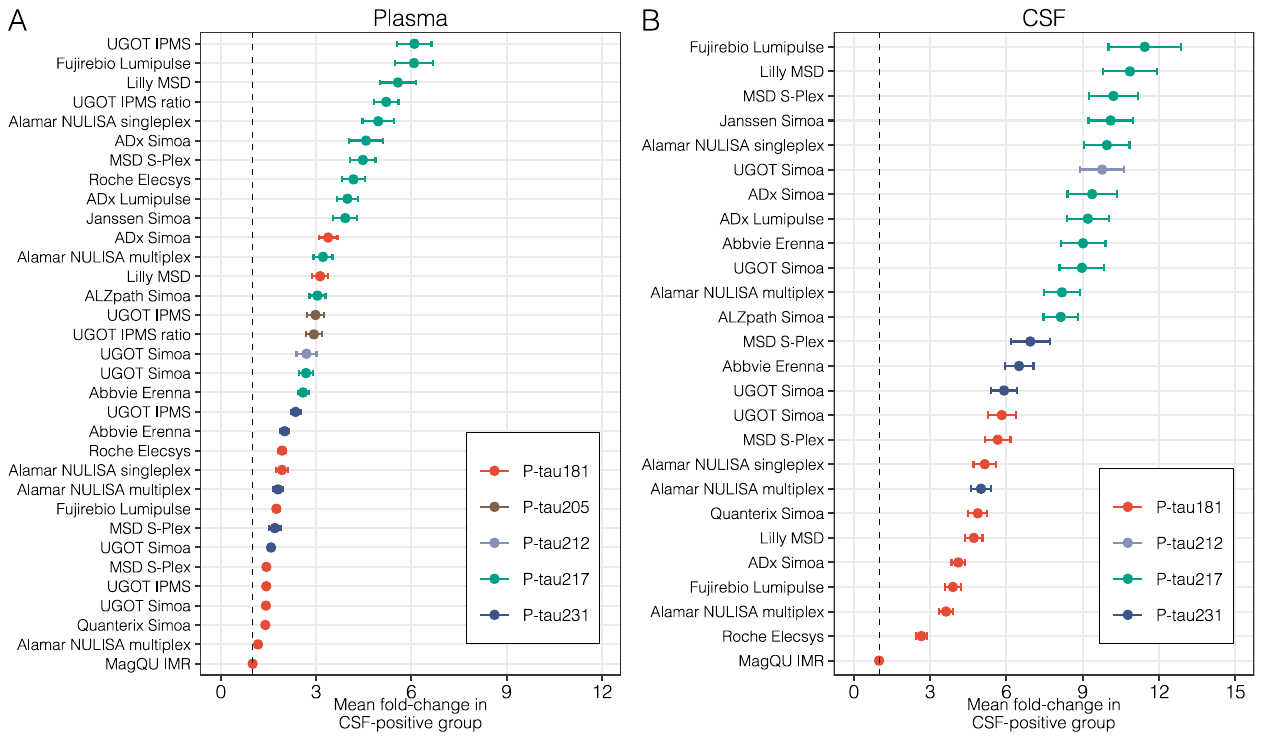

Forest plots indicate the mean fold-change of plasma (A) and cerebrospinal fluid (CSF; B) p-tau variants in the AD pathology group compared with the non-AD pathology group. Bars correspond to standard error. Supplementary Table 6 and Supplementary Table 7 numerically describe these plots.

**Supplementary Figure 5.** Discriminative ability of plasma p-tau assays to detect AD pathology.

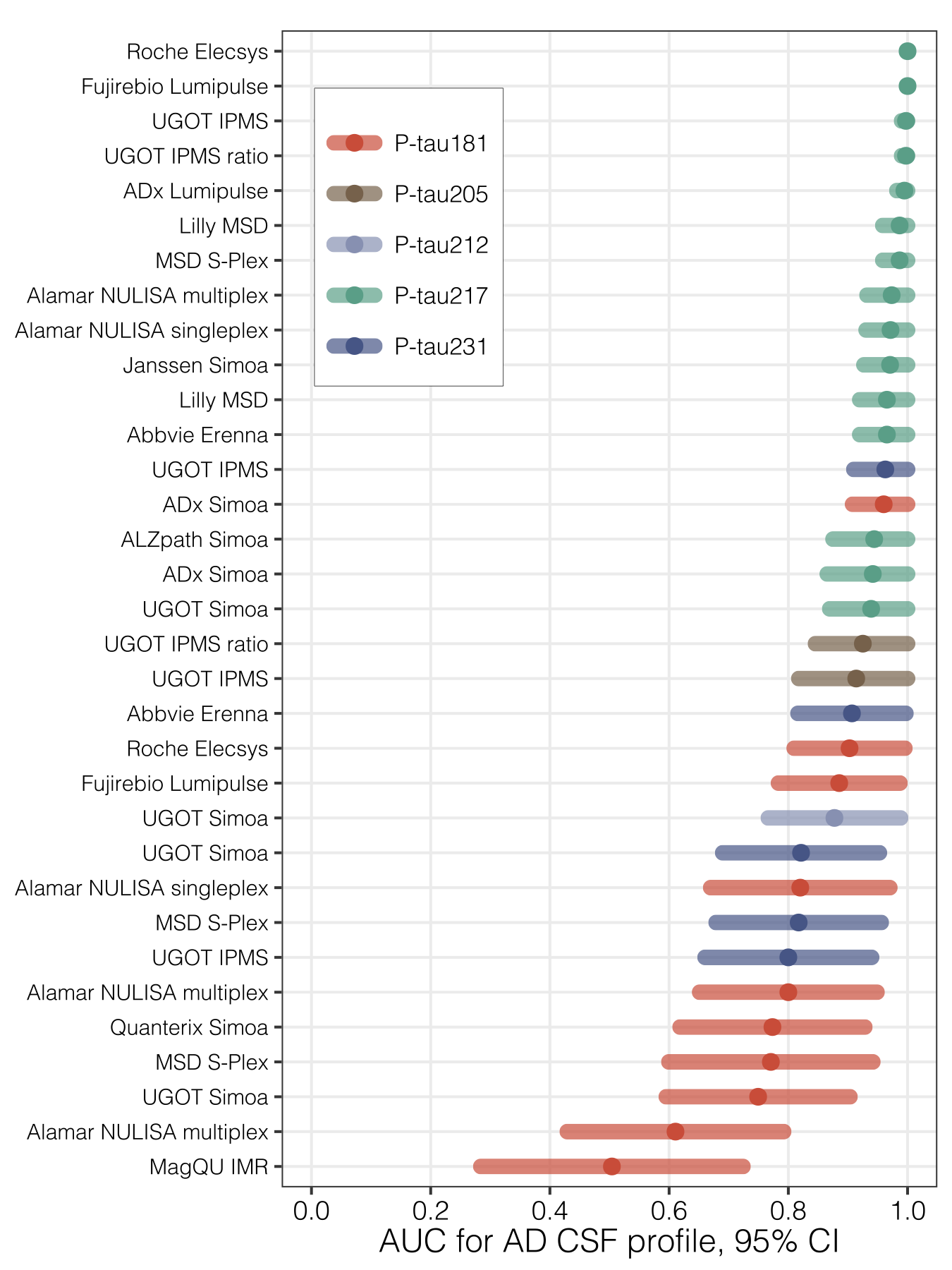

Forest plots displaying the discriminative ability of plasma p-tau assays to detect confirmed AD pathology (using an AD CSF profile as the reference standard), with the point estimate corresponding to the area under the receiver operating characteristics curve (AUC), and errorbars corresponding to 95% confidence intervals. These results are numerically described in Supplementary Table 8 and Supplementary Table 9.

**Supplementary Figure 6.** Intra-assay correlations between plasma and CSF p-tau181 biomarkers.

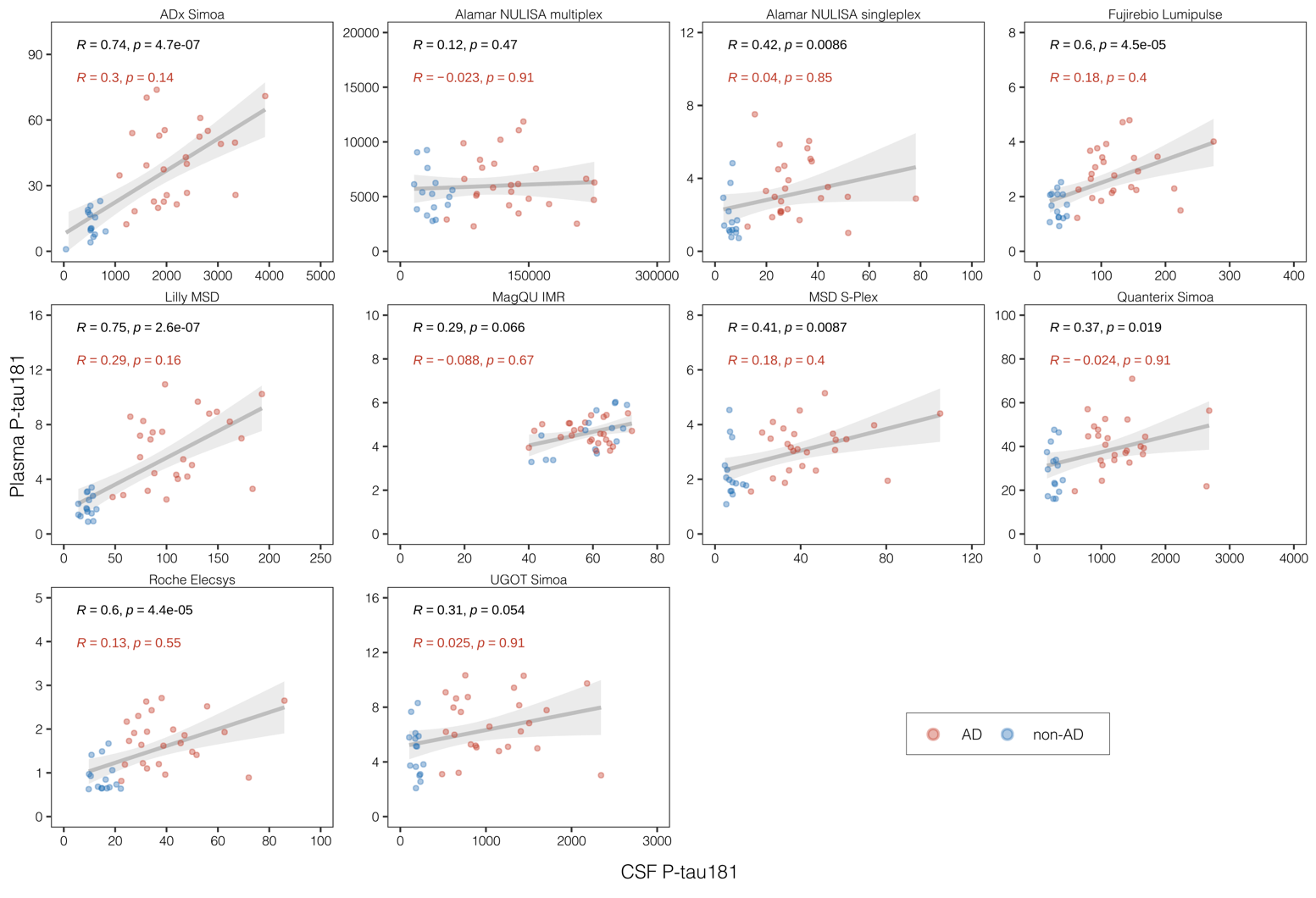

Scatterplots represent the associations between biomarker measurements in pg/mL performed with the same assay in plasma (y-axis) and cerebrospinal fluid (CSF; x-axis), alongside the mean regression line with 95% confidence intervals, computed based on data from all the participants in the cohort. Red dots indicate participants from the AD group and blue dots indicate participants from the non-AD group, as defined by clinical evaluation and CSF Aβ42/Aβ40 status. In each panel, the text in black indicates the Spearman’s correlation coefficient for the entire cohort, with red text indicating the Spearman’s correlation coefficient for the AD group only. *R* = Spearman’s Rho

**Supplementary Figure 7.** Intra-assay correlations between plasma and CSF p-tau231 biomarkers.

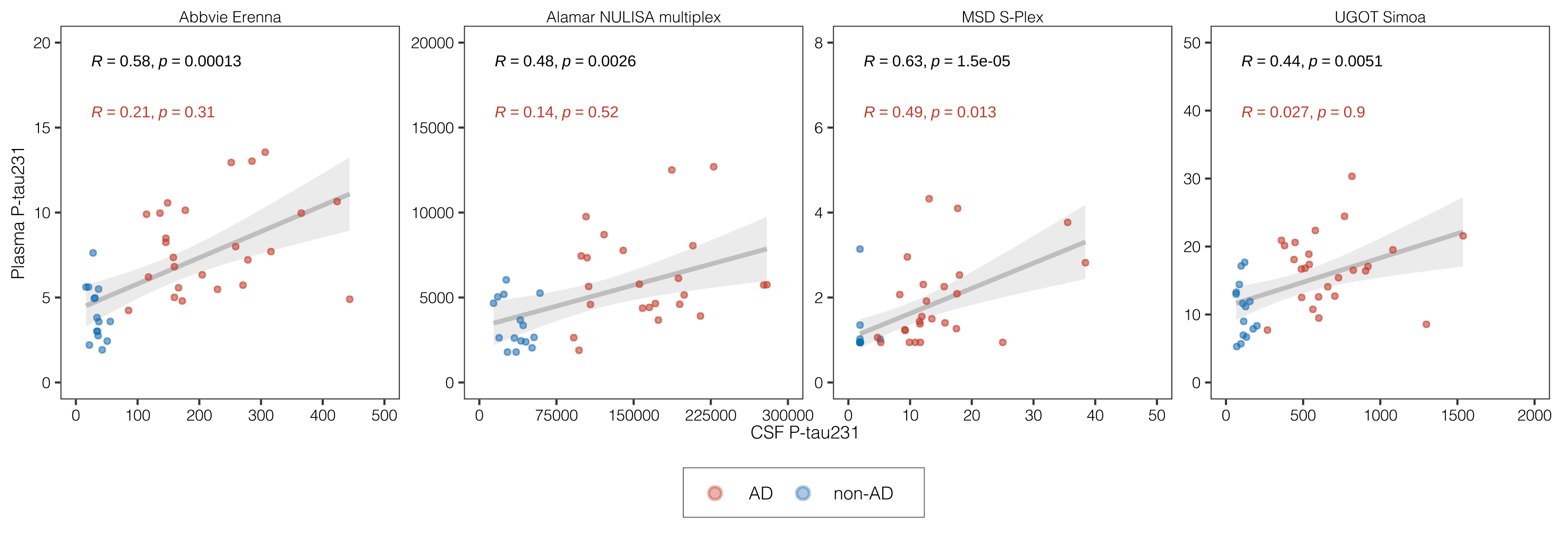

Scatterplots represent the associations between biomarker measurements in pg/mL performed with the same assay in plasma (y-axis) and cerebrospinal fluid (CSF; x-axis), alongside the mean regression line with 95% confidence intervals, computed based on data from all the participants in the cohort. Red dots indicate participants from the AD group and blue dots indicate participants from the non-AD group, as defined by clinical evaluation and CSF Aβ42/Aβ40 status. In each panel, the text in black indicates the Spearman’s correlation coefficient for the entire cohort and associated p value, with red text indicating these values for the AD group only. *R* = Spearman’s Rho

**Supplementary Figure 8.** Intra-assay correlation between plasma and CSF p-tau212.

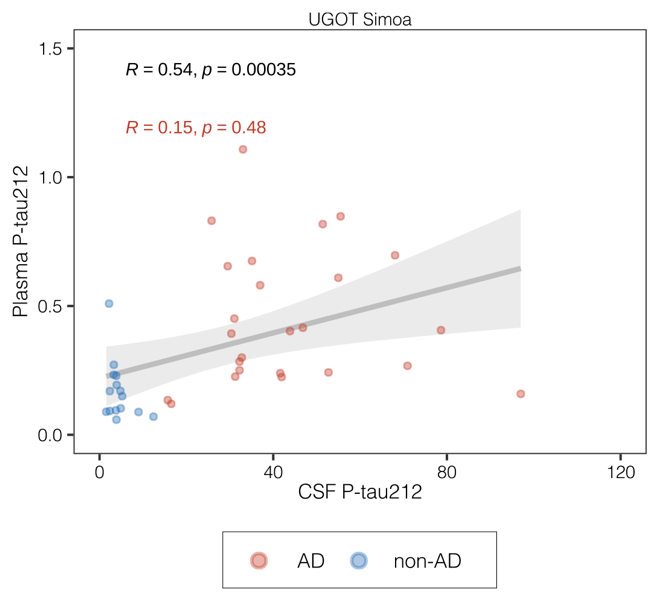

Scatterplot representing the association between biomarker measurements in pg/mL performed with the same p-tau212 assay in plasma (y-axis) and cerebrospinal fluid (CSF; x-axis), alongside the mean regression line with 95% confidence intervals, computed based on data from all the participants in the cohort. Red dots indicate participants from the AD group and blue dots indicate participants from the non-AD group, as defined by clinical evaluation and CSF Aβ42/Aβ40 status. The text in black indicates the Spearman’s correlation coefficient for the entire cohort and associated p value, with red text indicating these values for the AD group only *R* = Spearman’s Rho

**Supplementary Figure 9.** Associations between plasma p-tau assays and CSF LUMIPULSE G p-tau181.

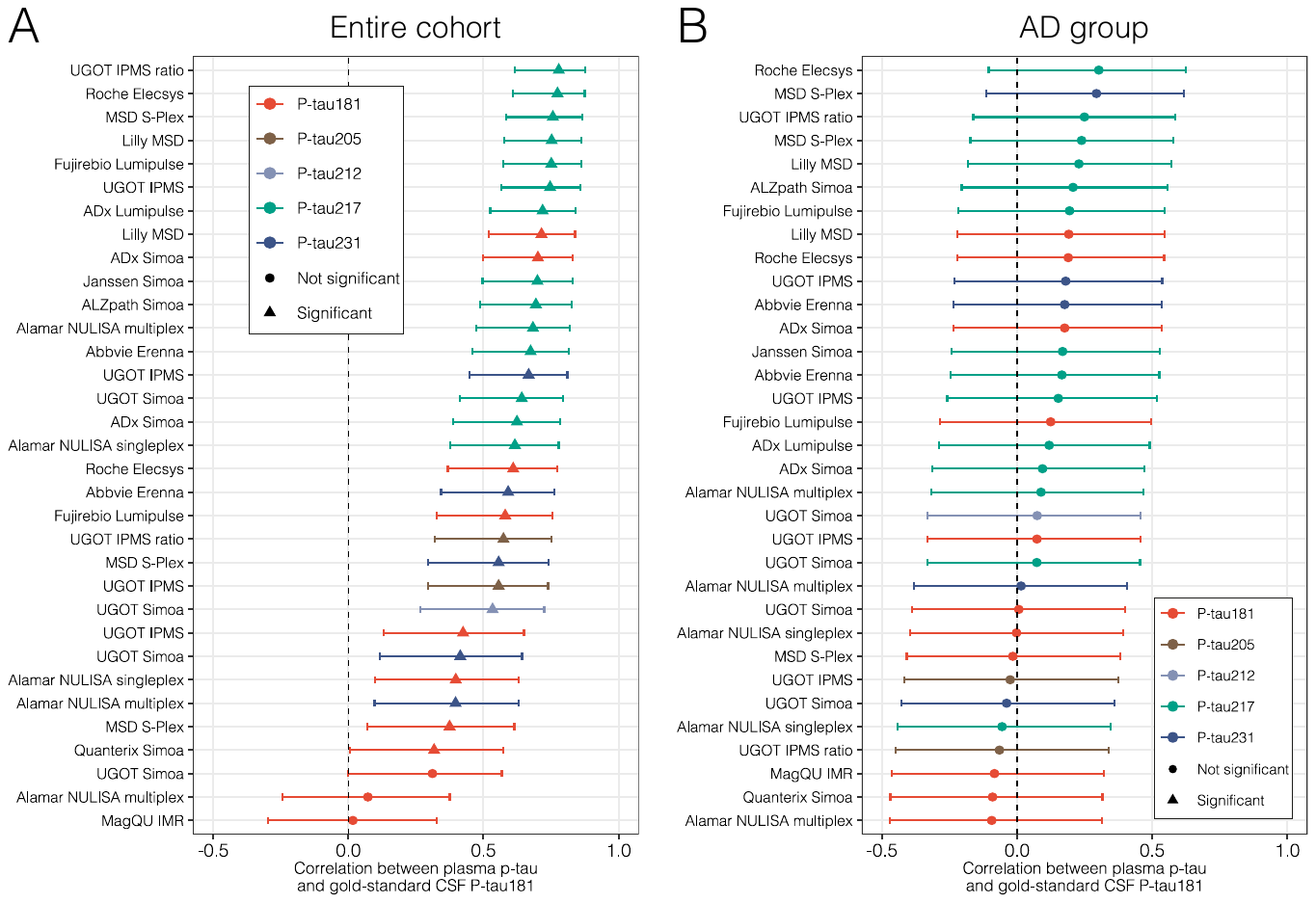

Forest plots displaying the Spearman correlation coefficients (with bootstrapped confidence intervals) between plasma p-tau assays and CSF LUMIPULSE G p-tau181, an FDA-approved gold standard metric for CSF p-tau. Associations are shown for the entire cohort (A) and in the AD group only (B).

**Supplementary Figure 10.** Passing-Bablok plots including candidate plasma p-tau217 reference materials.

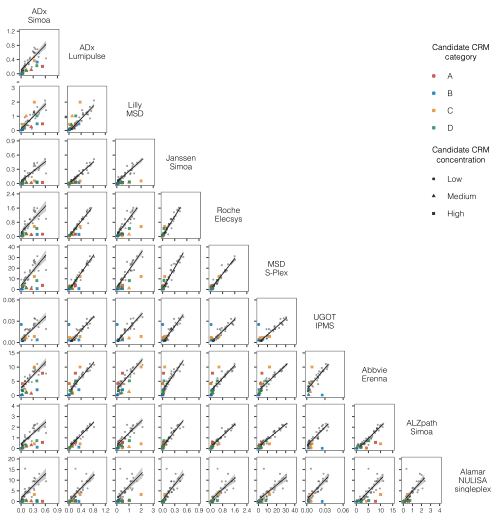

Scatterplots represent the continuous associations between all candidate reference material (CRM) for plasma p-tau217 assays. Four types of candidate CRM were evaluated (A, B, C, D) and each type is represented by the assigned colour. The shapes of individual data points indicate the candidate CRM concentrations. Participant samples are shown as grey circles.

A: full-length recombinant tau1–441 phosphorylated in vitro by glycogen synthase kinase 3β (TO8–50FN; SignalChem, Vancouver, BC, Canada) in Tau 2·0 Sample Diluent (Quanterix, #103847)

B: full-length recombinant tau1–441 in phosphate-buffered saline + 0·05% Tween

C: pooled EDTA plasma samples spiked with recombinant full-length tau

D: pooled EDTA plasma samples spiked with human CSF

**Supplementary Figure 11.** Bland-Altman plots including candidate p-tau217 reference materials.

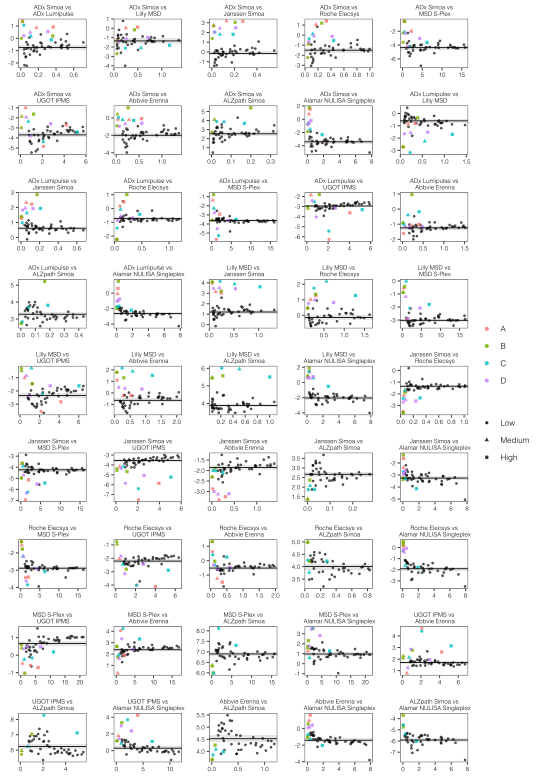

Bland-Altman plots comparing plasma p-tau217 assays for each candidate reference material (CRM). Four types of CRM were evaluated (A, B, C, D) and each type is represented by the assigned colour. The shapes of individual data points indicate the candidate CRM concentrations. Participant samples are shown as grey circles.

A: full-length recombinant tau1–441 phosphorylated in vitro by glycogen synthase kinase 3β in Tau 2·0 diluent

B: full-length recombinant tau1–441 in phosphate-buffered saline + 0·05% Tween

C: pooled EDTA plasma samples spiked with recombinant full-length tau

D: pooled EDTA plasma samples spiked with human CSF

**Supplementary Table 1.** CSF p-tau assay characteristics (listed if different from plasma assay in **Table 1**)

| **Participant** | **Target** | **Analytical platform** | **Functional LLOQ** | **LOD** | **Sample volume for duplicate (dead volume)** | **Sample dilution (sample diluent)** | **Calibrator** | **Calibrator range** | **Capture antibody** | **Detector antibody** | **Other assay details / Reference** |
| --- | --- | --- | --- | --- | --- | --- | --- | --- | --- | --- | --- |
| Abbvie | p-tau217 | Erenna | 0.15 pg/mL | 0.05 pg/mL | 4 µL (1 µL) | x50 (SMC Standard Diluent, Merck) | Full-length tau 441 expressed & phosphorylated in vivo by Sf9 cells | 0–36·45 pg/mL | ab288167 (epitope phosphorylated at T217) | Tau12 (N-terminal aa 6-18) | Method in supplement |
| Abbvie | p-tau231 | Erenna | 0.15 pg/mL | 0.05 pg/mL | 4 µL (1 µL) | x50 (SMC Standard Diluent, Merck) | Full-length tau 441 expressed & phosphorylated in vivo by Sf9 cells | 0–36·45 pg/mL | ab156624 (epitope phosphorylated at T231) | Tau12 (N-terminal aa 6-18) | Method in supplement |
| ADx NeuroSciences | p-tau181 | Simoa HD-X |  |  |  | x80 |  |  |  |  |  |
| ADx NeuroSciences | p-tau217 | Simoa HD-X |  |  |  | x20 |  |  |  |  |  |
| ADx NeuroSciences | p-tau217 | LUMIPULSE *G* 1200 |  |  |  | x1·45 |  |  |  |  |  |
| Alamar Biosciences, Inc | p-tau181 | NULISA qPCR (Singleplex)  NULISAseq (Multiplex) | 0.25 pg/mL  (Singleplex) | 0.015 pg/mL  (Singleplex) | 71 µL  (31 µL) | Neat sample with x5 (singleplex) or x10 (multiplex) onboard | Full-length recombinant tau 441 with site-specific phosphorylation at T181 | 0 – 920 pg/mL | Proprietary | Proprietary | Fully automated, NUcleic acid Linked Immuno-Sandwich Assay  Method in supplement |
| Alamar Biosciences, Inc | p-tau217 | NULISA qPCR (Singleplex)  NULISAseq (Multiplex) | 0.25 pg/mL  (Singleplex) | 0.019 pg/mL  (Singleplex) | 71 µL  (31 µL) | Neat sample with x5 (singleplex) or (multiplex) onboard | Full-length recombinant tau 441 with site-specific phosphorylation at T217 | 0 – 920 pg/mL | Proprietary | Proprietary | Fully automated, NUcleic acid Linked Immuno-Sandwich Assay  Method in supplement |
| Alamar Biosciences | p-tau231 | NULISA qPCR (Singleplex)  NULISAseq (Multiplex) | 0.25 pg/mL  (Singleplex) | 0.015pg/mL  (Singleplex) | 71 µL  (31 µL) | Neat sample with x5 (singleplex) or x10 (multiplex) onboard | Full-length recombinant tau 441 with site-specific phosphorylation at T231 | 0 – 920 pg/mL | Proprietary | Proprietary | Fully automated, NUcleic acid Linked Immuno-Sandwich Assay  Method in supplement |
| Janssen R&D | p-tau217 | Simoa HD-X | 0.088 pg/mL | 0.004 pg/ml | 200 µL (30 µL) | x8 (Simoa homebrew assay diluent) | Synthetic peptide (4.5kDa) = epitope of capture Ab-PEG4-epitope of detection Ab | 0-30 pg/ml | pT3 (epitope = 210-220, phosphorylated at T217, with enhanced binding with phosphorylation at T212) | hT43 (N-terminal aa 7-20) | 2-step HD-X setup (35-5). 75% helper beads. 50 ul RGP  Triana-Baltzer et al., 2020 |
| Fujirebio | p-tau181 | LUMIPULSE G | 0.282 pg/mL | 1·058 pg/mL | 40 µL (100 µL) | Not applicable | Same as plasma assay | 0-400 pg/mL | Same as plasma assay | Same as plasma assay | CE-IVDR Package Insert |
| Fujirebio | p-tau217 | LUMIPULSE G | Not available | Not available | 20 µL  (100 µL) | Not applicable | Same as plasma assay | 0-100 pg/mL | Same as plasma assay | Same as plasma assay | Not available, prototype used in this study |
| Lund University | p-tau217 | MSD Lilly | 0.46 pg/mL | 0.12 pg/mL | 30 µL (15 µL) | 1:4 (Low salt buffer) | Synthetic p-tau217 peptide | 0–180 pg/mL | Biotinylated-IBA493 (anti-p-tau217) | SULFO-TAG-4G10-E2 (Anti-tau) | Lund University |
| Lund University | p-tau181 | MSD Lilly | 1·62 pg/mL | 0.36 pg/mL | 30 µL (15 uL) | 1:4 (Low salt buffer) | Synthetic p-tau181 peptide | 0-200 pg/mL | Biotinylated-IBA406 (anti-p-tau181) | SULFO-TAG-4G10-E2 (Anti-tau) | Lund University |
| Meso Scale Diagnostics, LLC. (MSD) | p-tau181 | MSD S-PLEX |  |  | 25μL (5μL) | x2 (MSD Assay Diluent 2) |  |  |  |  | All other assay details are the same as plasma assay (Table 1). |
| Meso Scale Diagnostics, LLC. (MSD) | p-tau217 | MSD S-PLEX |  |  | 25μL (5μL) | x2 (MSD Assay Diluent 2) |  |  |  |  | All other assay details are the same as plasma assay (Table 1). |
| Meso Scale Diagnostics, LLC. (MSD) | p-tau231 | MSD S-PLEX |  |  | 25μL (5μL) | x2 (MSD Assay Diluent 2) |  |  |  |  | All other assay details are the same as plasma assay (Table 1). |
| Roche Diagnostics | p-tau181 | Cobas e (Elecsys) | 8 pg/mL | 8 pg/mL | 30 uL (approx. 100 µL) singlicate | No dilution required | Synthetic peptide | 8-120 pg/mL | 11H5V1 | PC1C6 | Electrochemiluminescence sandwich immunoassay, 18 min total incubation time  Lifke et al., 2019 ^1^ |
| University of Gothenburg (UGOT) | p-tau181 | Simoa HD-X |  |  |  | x20 |  |  |  |  |  |
| University of Gothenburg (UGOT) | p-tau212 | Simoa HD-X | 0.073 pg/mL | 0.01 pg/mL | 240 µL (40 µL) | x10 (Tau 2·0, Quanterix) | Full-length recombinant tau 441 phosphorylated in vitro by DYRK1A (Abcam269022) | 0–41·67 pg/mL | p-Tau212·7B3 (epitope p-Tau212) | Tau12 (N-terminal aa 6-18) | 2-step HD-X set-up (47-7). 0% helper beads.  Kac et al., 2023 |
| University of Gothenburg (UGOT) | p-tau217 | Simoa HD-X | 0.08 pg/mL | 0.01 pg/mL | 200 µL | x20 | GSK | 1-64 pg/mL | p.T217.FG | Tau12 | University of Gothenburg (UGOT) |
| University of Gothenburg (UGOT) | p-tau231 | Simoa HD-X |  |  |  | x20 |  |  |  |  |  |

**Supplementary Table 2.** Analytical performance of plasma and CSF assays

| **Biomarker** | **Assay** | **Matrix** | **Internal Quality Controls** | | | **Samples** | | |
| --- | --- | --- | --- | --- | --- | --- | --- | --- |
|  |  |  | **QC Description** | **Repeatability (%)** | **Intermediate precision (%)** | **Repeatability (%)** | **No. missing samples** | **No. missing replicates** |
| p-tau181 | ADx Simoa | Plasma | EDTA plasma QC panel (n = 3) | < 20% | < 20% | Mean %CV [95% CI]: 12·73 [6·65 to 18·8]  Min: 0·15 to Max 80·1 | 1 (AEB below LLOQ) | 2 |
| p-tau181 | ADx Simoa | CSF | RV samples (n = 2) | < 10% | < 15% | Mean %CV [95% CI]: 8·34 [5·26 to 11·42]  Min: 0·1 to Max 55·84 | 0 | 0 |
| p-tau181 | Alamar NULISA multiplex | Plasma | CSF-spiked EDTA plasma | 4·9% | NA (all samples in 1 run) | NA (single rep) | 0 | NA |
| p-tau181 | Alamar NULISA multiplex | CSF | CSF-spiked EDTA plasma | 4·9% | NA (all samples in 1 run) | NA (single rep) | 0 | NA |
| p-tau181 | Alamar NULISA singleplex | Plasma | Calibrator-spiked EDTA plasma at 2 levels | 8·0% | 8·7% | 19·0% | 0 | 0 |
| p-tau181 | Alamar NULISA singleplex | CSF | Calibrator-spiked EDTA plasma at 2 levels | 8·0% | 8·7% | 5·4% | 0 | 0 |
| p-tau181 | Fujirebio Lumipulse | Plasma | Buffered matrix with added synthetic peptide, 2 levels |  |  |  |  |  |
| p-tau181 | Lilly MSD | Plasma | 3 pooled plasma QCs | 5·0% | 7·4% | 4·6% | N/A | N/A |
| p-tau181 | Lilly MSD | CSF | 2 pooled CSF QCs | 7·5% | 11% | 3·2% | N/A | N/A |
| p-tau181 | MagQU IMR^a^ | Plasma |  |  |  |  |  |  |
| p-tau181 | MagQU IMR^a^ | CSF |  |  |  |  |  |  |
| p-tau181 | MSD S-Plex | Plasma | Calibrator-spiked assay diluent at three levels | 1·7 | 4·0 | 4·4 | 0 | 0 |
| p-tau181 | MSD S-Plex | CSF | Calibrator-spiked assay diluent at three levels | 1·7 | 4·0 | 3·7 | 0 | 0 |
| p-tau181 | Quanterix Simoa^a^ | Plasma |  |  |  |  |  |  |
| p-tau181 | Quanterix Simoa^a^ | CSF |  |  |  |  |  |  |
| p-tau181 | Roche Elecsys | Plasma | Proprietary, 2 levels | ≤3·9% | N/A | ≤6·1% | 0 | 0 |
| p-tau181 | Roche Elecsys | CSF | Buffered matrix with added synthetic peptide, 2 levels | ≤1·5% | N/A | ≤2·4% | 0 | 0 |
| p-tau181 | UGOT IPMS^a^ | Plasma |  |  |  |  |  |  |
| p-tau181 | UGOT Simoa | Plasma | Pooled plasma | 5·2% | N/A | 7·4 | 0 | 2 |
| p-tau181 | UGOT Simoa | CSF | Pooled plasma | 4·4% | N/A | 7·7 | 0 | 0 |
| p-tau205 | UGOT IPMS^a^ | Plasma |  |  |  |  |  |  |
| p-tau205 | UGOT IPMS ratio^a^ | Plasma |  |  |  |  |  |  |
| p-tau212 | UGOT Simoa | Plasma | Plasma pool | 2·6-7·6% | N/A | 8 | 0 | 1 |
| p-tau212 | UGOT Simoa | CSF | CSF pool | 0.9-8·7% | N/A | 4·3 | 0 | 0 |
| p-tau217 | Abbvie Erenna^a^ | Plasma |  |  |  |  |  |  |
| p-tau217 | Abbvie Erenna^a^ | CSF |  |  |  |  |  |  |
| p-tau217 | ADx Lumipulse | Plasma | EDTA plasma QC panel (n = 3) | < 10% | < 15% | NA | 0 | NA |
| p-tau217 | ADx Lumipulse | CSF | EDTA plasma QC panel (n = 3) | < 10% | < 15% | NA | 0 | NA |
| p-tau217 | ADx Simoa | Plasma | EDTA plasma QC panel (n = 3) | < 20% | < 20% | Mean %CV [95% CI]: 17·53 [9·3 – 25,75]  Min: 0·3 - Max 121·7 | 4 (AEB below LLOQ) | 3 |
| p-tau217 | ADx Simoa | CSF | RV samples (n = 2) | < 10% | < 15% | Mean %CV [95% CI]: 6·91 [5,08 – 9·09]  Min: 0·21 - Max 30 | 0 | 1 |
| p-tau217 | Alamar NULISA multiplex | Plasma | CSF-spiked EDTA plasma | 9·3% | NA (all samples in 1 run) | NA (single rep) | 0 | NA |
| p-tau217 | Alamar NULISA multiplex | CSF | CSF-spiked EDTA plasma | 9·3% | NA (all samples in 1 run) | NA (single rep) | 0 | NA |
| p-tau217 | Alamar NULISA singleplex | Plasma | Calibrator-spiked EDTA plasma at 2 levels | 6·4% | 6·9% | 8·9% | 0 | 0 |
| p-tau217 | Alamar NULISA singleplex | CSF | Calibrator-spiked EDTA plasma at 2 levels | 6·4% | 6·9% | 7·8% | 0 | 0 |
| p-tau217 | Fujirebio Lumipulse | Plasma | Buffered matrix with added synthetic peptide, 2 levels |  |  |  |  |  |
| p-tau217 | Fujirebio Lumipulse | CSF | Buffered matrix with added synthetic peptide, 2 levels |  |  |  |  |  |
| p-tau217 | Janssen Simoa | Plasma | 3 peptide/buffer QCs in each run, 7 plasma pools in one run | 4·08 | 4·93 | 4·43 | 0 | 0 |
| p-tau217 | Janssen Simoa | CSF | 3 peptide/buffer QCs in each run, | 6·40 | 9·16 | 3·79 | 0 | 0 |
| p-tau217 | Lilly MSD | Plasma | 3 pooled plasma QCs | 5·6% | 3·6% | 6·5% | N/A | N/A |
| p-tau217 | Lilly MSD | CSF | 2 pooled CSF QCs | 2·1% | 3·2% | 2·2% | N/A | N/A |
| p-tau217 | MSD S-Plex | Plasma | Calibrator-spiked assay diluent at three levels | 1·7 | 3·0 | 2·5 | 0 | 0 |
| p-tau217 | MSD S-Plex | CSF | Calibrator-spiked assay diluent at three levels | 1·7 | 3·0 | 3·2 | 0 | 0 |
| p-tau217 | Roche Elecsys | Plasma | N/A | N/A | N/A | N/A (all samples run in singlicate) | 2 | N/A |
| p-tau217 | UGOT IPMS^a^ | Plasma |  |  |  |  |  |  |
| p-tau217 | UGOT IPMS ratio^a^ | Plasma |  |  |  |  |  |  |
| p-tau217 | UGOT Simoa | Plasma | Pooled plasma | 3·2% | N/A | 8·3 | 1 | 4 |
| p-tau217 | UGOT Simoa | CSF | Pooled plasma | 3·4% | N/A | 10·7 | No | 1 |
| p-tau231 | Abbvie Erenna^a^ | Plasma |  |  |  |  |  |  |
| p-tau231 | Abbvie Erenna^a^ | CSF |  |  |  |  |  |  |
| p-tau231 | Alamar NULISA multiplex | Plasma | CSF-spiked EDTA plasma | 3·3% | NA (all samples in 1 run) | NA (single rep) | 0 | NA |
| p-tau231 | Alamar NULISA multiplex | CSF | CSF-spiked EDTA plasma | 3·3% | NA (all samples in 1 run) | NA (single rep) | 0 | NA |
| p-tau231 | MSD S-Plex | Plasma | Calibrator-spiked assay diluent at three levels | 3·4 | 6·2 | 15·8 | 16 | 6 |
| p-tau231 | MSD S-Plex | CSF | Calibrator-spiked assay diluent at three levels | 3·4 | 6·2 | 5·4 | 13 | 1 |
| p-tau231 | UGOT Simoa | Plasma | Pooled plasma | 2·4% | N/A | 5·3 | 2 | 2 |
| p-tau231 | UGOT Simoa | CSF | Pooled plasma | 5·4% | N/A | 11·7 | 0 | 0 |

^a^ Data not provided by vendor

**Supplementary Table 3.** Description of Candidate Certified Reference Materials

| **Candidate CRM** | **Component 1** | **Component 2** | **Calculated average enzyme per bead (AEB)** |
| --- | --- | --- | --- |
| A-High | Full-length recombinant tau phosphorylated in vitro by GSK 3β | Tau 2·0 Sample Diluent | 0·898 |
| A-Medium |  |  | 0·699 |
| A-Low |  |  | 0·135 |
| B-High | Full-length recombinant tau phosphorylated in vitro by GSK 3β | PBS + 0·05% Tween | 0·728 |
| B-Medium |  |  | 0·341 |
| B-Low |  |  | 0·107 |
| C-High | Human EDTA plasma pool | Tau 2·0 Sample Diluent | 0·824 |
| C-Medium |  |  | 0·388 |
| C-Low |  |  | 0·182 |
| D-High | Human EDTA plasma pool | Human CSF; PBS + 0·05% Tween | 1·02 |
| D-Medium |  |  | 0·415 |
| D-Low |  |  | 0·197 |

**Supplementary Table 4.** Median fold-change in AD pathology group for plasma and CSF p-tau217 biomarker assays.

| **Biomarker** | **Assay** | **Matrix** | **Fold change (SE)** |
| --- | --- | --- | --- |
| p-tau217 | Abbvie Erenna | Plasma | 2·65 (0·17) |
| p-tau217 | Abbvie Erenna | CSF | 6·99 (0·88) |
| p-tau217 | ADx Lumipulse | Plasma | 3·88 (0·34) |
| p-tau217 | ADx Lumipulse | CSF | 7·53 (0·83) |
| p-tau217 | ADx Simoa | Plasma | 4·32 (0·54) |
| p-tau217 | ADx Simoa | CSF | 8·3 (0·97) |
| p-tau217 | Alamar NULISA multiplex | Plasma | 2·93 (0·3) |
| p-tau217 | Alamar NULISA multiplex | CSF | 8·24 (0·7) |
| p-tau217 | Alamar NULISA singleplex | Plasma | 4·3 (0·49) |
| p-tau217 | Alamar NULISA singleplex | CSF | 8·35 (0·89) |
| p-tau217 | ALZpath Simoa | Plasma | 3·18 (0·25) |
| p-tau217 | ALZpath Simoa | CSF | 7·07 (0·68) |
| p-tau217 | Fujirebio Lumipulse | Plasma | 5·69 (0·61) |
| p-tau217 | Fujirebio Lumipulse | CSF | 9·73 (1·42) |
| p-tau217 | Janssen Simoa | Plasma | 3·5 (0·38) |
| p-tau217 | Janssen Simoa | CSF | 9·17 (0·87) |
| p-tau217 | Lilly MSD | Plasma | 5·21 (0·53) |
| p-tau217 | Lilly MSD | CSF | 9·24 (1·07) |
| p-tau217 | MSD S-Plex | Plasma | 4·49 (0·4) |
| p-tau217 | MSD S-Plex | CSF | 8·74 (0·97) |
| p-tau217 | Roche Elecsys | Plasma | 4·09 (0·37) |
| p-tau217 | Roche Elecsys | CSF | N/A |
| p-tau217 | UGOT IPMS | Plasma | 5·8 (0·54) |
| p-tau217 | UGOT IPMS | CSF | N/A |
| p-tau217 | UGOT IPMS ratio | Plasma | 5·19 (0·39) |
| p-tau217 | UGOT IPMS ratio | CSF | N/A |
| p-tau217 | UGOT Simoa | Plasma | 2·56 (0·23) |
| p-tau217 | UGOT Simoa | CSF | 7·89 (0·88) |

**Supplementary Table 5.** Median fold-change in AD pathology group for plasma and CSF p-tau181, p-tau205, p-tau212 and p-tau231 biomarkers.

| **Biomarker** | **Assay** | **Matrix** | **Fold change (SE)** |
| --- | --- | --- | --- |
| P-tau181 | ADx Simoa | Plasma | 3·26 (0·29) |
| P-tau181 | ADx Simoa | CSF | 3·7 (0·27) |
| P-tau181 | Alamar NULISA multiplex | Plasma | 1·13 (0·1) |
| P-tau181 | Alamar NULISA multiplex | CSF | 3·56 (0·27) |
| P-tau181 | Alamar NULISA singleplex | Plasma | 1·64 (0·18) |
| P-tau181 | Alamar NULISA singleplex | CSF | 4·37 (0·44) |
| P-tau181 | Fujirebio Lumipulse | Plasma | 1·7 (0·11) |
| P-tau181 | Fujirebio Lumipulse | CSF | 3·54 (0·32) |
| P-tau181 | Lilly MSD | Plasma | 3·43 (0·25) |
| P-tau181 | Lilly MSD | CSF | 4·29 (0·34) |
| P-tau181 | MagQU IMR | Plasma | 0·99 (0·02) |
| P-tau181 | MagQU IMR | CSF | 1·01 (0·03) |
| P-tau181 | MSD S-Plex | Plasma | 1·46 (0·08) |
| P-tau181 | MSD S-Plex | CSF | 4·91 (0·51) |
| P-tau181 | Quanterix Simoa | Plasma | 1·36 (0·08) |
| P-tau181 | Quanterix Simoa | CSF | 4·49 (0·38) |
| P-tau181 | Roche Elecsys | Plasma | 1·9 (0·13) |
| P-tau181 | Roche Elecsys | CSF | 2·43 (0·21) |
| P-tau181 | UGOT IPMS | Plasma | 1·44 (0·08) |
| P-tau181 | UGOT IPMS | CSF | N/A |
| P-tau181 | UGOT Simoa | Plasma | 1·38 (0·09) |
| P-tau181 | UGOT Simoa | CSF | 4·74 (0·54) |
| P-tau205 | UGOT IPMS | Plasma | 3·03 (0·27) |
| P-tau205 | UGOT IPMS | CSF | N/A |
| P-tau205 | UGOT IPMS ratio | Plasma | 2·86 (0·24) |
| P-tau205 | UGOT IPMS ratio | CSF | N/A |
| P-tau212 | UGOT Simoa | Plasma | 2·39 (0·31) |
| P-tau212 | UGOT Simoa | CSF | 8·31 (0·87) |
| P-tau231 | Abbvie Erenna | Plasma | 1·91 (0·14) |
| P-tau231 | Abbvie Erenna | CSF | 5·22 (0·57) |
| P-tau231 | Alamar NULISA multiplex | Plasma | 1·67 (0·16) |
| P-tau231 | Alamar NULISA multiplex | CSF | 4·85 (0·39) |
| P-tau231 | MSD S-Plex | Plasma | 1·33 (0·18) |
| P-tau231 | MSD S-Plex | CSF | 5·76 (0·76) |
| P-tau231 | UGOT IPMS | Plasma | 2·29 (0·15) |
| P-tau231 | UGOT IPMS | CSF | N/A |
| P-tau231 | UGOT Simoa | Plasma | 1·58 (0·1) |
| P-tau231 | UGOT Simoa | CSF | 5·2 (0·51) |

**Supplementary Table 6.** Mean fold-change in AD pathology group individuals for plasma

and CSF p-tau217 biomarkers.

| **Biomarker** | **Assay** | **Matrix** | **Fold change (SE)** |
| --- | --- | --- | --- |
| P-tau217 | Abbvie Erenna | Plasma | 2·59 (0·17) |
| P-tau217 | Abbvie Erenna | CSF | 9·02 (0·88) |
| P-tau217 | ADx Lumipulse | Plasma | 3·99 (0·34) |
| P-tau217 | ADx Lumipulse | CSF | 9·2 (0·83) |
| P-tau217 | ADx Simoa | Plasma | 4·57 (0·54) |
| P-tau217 | ADx Simoa | CSF | 9·37 (0·97) |
| P-tau217 | Alamar NULISA multiplex | Plasma | 3·22 (0·3) |
| P-tau217 | Alamar NULISA multiplex | CSF | 8·19 (0·7) |
| P-tau217 | Alamar NULISA singleplex | Plasma | 4·96 (0·49) |
| P-tau217 | Alamar NULISA singleplex | CSF | 9·95 (0·89) |
| P-tau217 | ALZpath Simoa | Plasma | 3·04 (0·25) |
| P-tau217 | ALZpath Simoa | CSF | 8·14 (0·68) |
| P-tau217 | Fujirebio Lumipulse | Plasma | 6·08 (0·61) |
| P-tau217 | Fujirebio Lumipulse | CSF | 11·44 (1·42) |
| P-tau217 | Janssen Simoa | Plasma | 3·92 (0·38) |
| P-tau217 | Janssen Simoa | CSF | 10·1 (0·87) |
| P-tau217 | Lilly MSD | Plasma | 5·30 (0·53) |
| P-tau217 | Lilly MSD | CSF | 10·86 (1·07) |
| P-tau217 | MSD S-Plex | Plasma | 4·47 (0·4) |
| P-tau217 | MSD S-Plex | CSF | 10·21 (0·97) |
| P-tau217 | Roche Elecsys | Plasma | 4·18 (0·37) |
| P-tau217 | UGOT IPMS | Plasma | 6·1 (0·54) |
| P-tau217 | UGOT IPMS | CSF | N/A |
| P-tau217 | UGOT IPMS ratio | Plasma | 5·21 (0·39) |
| P-tau217 | UGOT IPMS ratio | CSF | N/A |
| P-tau217 | UGOT Simoa | Plasma | 2·68 (0·23) |
| P-tau217 | UGOT Simoa | CSF | 8·97 (0·88) |

**Supplementary Table 7.** Mean fold-change in AD pathology group for plasma and CSF p-tau181, p-tau205, p-tau212 and p-tau231 biomarkers.

| **Biomarker** | **Assay** | **Matrix** | **Fold change (SE)** |
| --- | --- | --- | --- |
| P-tau181 | ADx Simoa | Plasma | 3·38 (0·29) |
| P-tau181 | ADx Simoa | CSF | 4·11 (0·27) |
| P-tau181 | Alamar NULISA multiplex | Plasma | 1·17 (0·1) |
| P-tau181 | Alamar NULISA multiplex | CSF | 3·62 (0·27) |
| P-tau181 | Alamar NULISA singleplex | Plasma | 1·92 (0·18) |
| P-tau181 | Alamar NULISA singleplex | CSF | 5·15 (0·44) |
| P-tau181 | Fujirebio Lumipulse | Plasma | 1·75 (0·11) |
| P-tau181 | Fujirebio Lumipulse | CSF | 3·89 (0·32) |
| P-tau181 | Lilly MSD | Plasma | 3·13 (0·25) |
| P-tau181 | Lilly MSD | CSF | 4·72 (0·34) |
| P-tau181 | MagQU IMR | Plasma | 1 (0·02) |
| P-tau181 | MagQU IMR | CSF | 0·99 (0·03) |
| P-tau181 | MSD S-Plex | Plasma | 1·43 (0·08) |
| P-tau181 | MSD S-Plex | CSF | 5·66 (0·51) |
| P-tau181 | Quanterix Simoa | Plasma | 1·4 (0·08) |
| P-tau181 | Quanterix Simoa | CSF | 4·87 (0·38) |
| P-tau181 | Roche Elecsys | Plasma | 1·93 (0·13) |
| P-tau181 | Roche Elecsys | CSF | 2·65 (0·21) |
| P-tau181 | UGOT IPMS | Plasma | 1·43 (0·08) |
| P-tau181 | UGOT IPMS | CSF | N/A |
| P-tau181 | UGOT Simoa | Plasma | 1·42 (0·09) |
| P-tau181 | UGOT Simoa | CSF | 5·82 (0·54) |
| P-tau205 | UGOT IPMS | Plasma | 2·98 (0·27) |
| P-tau205 | UGOT IPMS | CSF | N/A |
| P-tau205 | UGOT IPMS ratio | Plasma | 2·93 (0·24) |
| P-tau205 | UGOT IPMS ratio | CSF | N/A |
| P-tau212 | UGOT Simoa | Plasma | 2·7 (0·31) |
| P-tau212 | UGOT Simoa | CSF | 9·77 (0·87) |
| P-tau231 | Abbvie Erenna | Plasma | 2·59 (0·17) |
| P-tau231 | Abbvie Erenna | CSF | 9·02 (0·88) |
| P-tau231 | Alamar NULISA multiplex | Plasma | 3·99 (0·34) |
| P-tau231 | Alamar NULISA multiplex | CSF | 9·2 (0·83) |
| P-tau231 | MSD S-Plex | Plasma | 4·57 (0·54) |
| P-tau231 | MSD S-Plex | CSF | 9·37 (0·97) |
| P-tau231 | UGOT IPMS | Plasma | 3·22 (0·3) |
| P-tau231 | UGOT IPMS | CSF | N/A |
| P-tau231 | UGOT Simoa | Plasma | 8·19 (0·7) |
| P-tau231 | UGOT Simoa | CSF | 4·96 (0·49) |

**Supplementary Table 8**. AUCs for detecting CSF AD pathology for plasma and CSF p-tau217 biomarkers.

| **Biomarker** | **Assay** | **Matrix** | **AUC (95% CI)** |
| --- | --- | --- | --- |
| P-tau217 | Abbvie Erenna | Plasma | 0·97 (0·92–1) |
| P-tau217 | Abbvie Erenna | CSF | 1 (1–1) |
| P-tau217 | ADx Lumipulse | Plasma | 0·99 (0·98–1) |
| P-tau217 | ADx Lumipulse | CSF | 1 (1–1) |
| P-tau217 | ADx Simoa | Plasma | 0·94 (0·87–1) |
| P-tau217 | ADx Simoa | CSF | 1 (1–1) |
| P-tau217 | Alamar NULISA multiplex | Plasma | 0·97 (0·93–1) |
| P-tau217 | Alamar NULISA multiplex | CSF | 1 (1–1) |
| P-tau217 | Alamar NULISA singleplex | Plasma | 0·97 (0·93–1) |
| P-tau217 | Alamar NULISA singleplex | CSF | 1 (1–1) |
| P-tau217 | ALZpath Simoa | Plasma | 0·94 (0·87–1) |
| P-tau217 | ALZpath Simoa | CSF | 1 (1–1) |
| P-tau217 | Fujirebio Lumipulse | Plasma | 1 (1–1) |
| P-tau217 | Fujirebio Lumipulse | CSF | 1 (0·99–1) |
| P-tau217 | Janssen Simoa | Plasma | 0·97 (0·93–1) |
| P-tau217 | Janssen Simoa | CSF | 1 (1–1) |
| P-tau217 | Lilly MSD | Plasma | 0·99 (0·96–1) |
| P-tau217 | Lilly MSD | CSF | 1 (1–1) |
| P-tau217 | MSD S-Plex | Plasma | 0·99 (0·96–1) |
| P-tau217 | MSD S-Plex | CSF | 1 (1–1) |
| P-tau217 | Roche Elecsys | Plasma | 0·99 (0·98–1) |
| P-tau217 | UGOT IPMS | Plasma | 1 (0·99–1) |
| P-tau217 | UGOT IPMS | CSF | N/A |
| P-tau217 | UGOT IPMS ratio | Plasma | 1 (0·99–1) |
| P-tau217 | UGOT IPMS ratio | CSF | N/A |
| P-tau217 | UGOT Simoa | Plasma | 0·94 (0·87–1) |
| P-tau217 | UGOT Simoa | CSF | 1 (1–1) |

**Supplementary Table 9**. AUCs for detecting CSF AD pathology for plasma and CSF p-tau181, p-tau205, p-tau212 and p-tau231 biomarkers.

| **Biomarker** | **Assay** | **Matrix** | **AUC (95% CI)** |
| --- | --- | --- | --- |
| P-tau181 | ADx Simoa | Plasma | 0·96 (0·91–1) |
| P-tau181 | ADx Simoa | CSF | 1 (1–1) |
| P-tau181 | Alamar NULISA multiplex | Plasma | 0·61 (0·43–0·79) |
| P-tau181 | Alamar NULISA multiplex | CSF | 0·99 (0·97–1) |
| P-tau181 | Alamar NULISA singleplex | Plasma | 0·82 (0·67–0·97) |
| P-tau181 | Alamar NULISA singleplex | CSF | 1 (1–1) |
| P-tau181 | Fujirebio Lumipulse | Plasma | 0·89 (0·78–0·99) |
| P-tau181 | Fujirebio Lumipulse | CSF | 1 (1–1) |
| P-tau181 | Lilly MSD | Plasma | 0·97 (0·92–1) |
| P-tau181 | Lilly MSD | CSF | 1 (1–1) |
| P-tau181 | MagQU IMR | Plasma | 0·5 (0·28–0·72) |
| P-tau181 | MagQU IMR | CSF | 0·55 (0·35–0·75) |
| P-tau181 | MSD S-Plex | Plasma | 0·77 (0·6–0·94) |
| P-tau181 | MSD S-Plex | CSF | 1 (1–1) |
| P-tau181 | Quanterix Simoa | Plasma | 0·77 (0·62–0·93) |
| P-tau181 | Quanterix Simoa | CSF | 1 (1–1) |
| P-tau181 | Roche Elecsys | Plasma | 0·9 (0·81–1) |
| P-tau181 | Roche Elecsys | CSF | 1 (1–1) |
| P-tau181 | UGOT IPMS | Plasma | 0·8 (0·65–0·95) |
| P-tau181 | UGOT IPMS | CSF | N/A |
| P-tau181 | UGOT Simoa | Plasma | 0·75 (0·6–0·9) |
| P-tau181 | UGOT Simoa | CSF | 1 (1–1) |
| P-tau205 | UGOT IPMS | Plasma | 0·91 (0·82–1) |
| P-tau205 | UGOT IPMS | CSF | N/A |
| P-tau205 | UGOT IPMS ratio | Plasma | 0·93 (0·85–1) |
| P-tau205 | UGOT IPMS ratio | CSF | N/A |
| P-tau212 | UGOT Simoa | Plasma | 0·88 (0·77–0·99) |
| P-tau212 | UGOT Simoa | CSF | 1 (1–1) |
| P-tau231 | Abbvie Erenna | Plasma | 0·91 (0·82–1) |
| P-tau231 | Abbvie Erenna | CSF | 1 (1–1) |
| P-tau231 | Alamar NULISA multiplex | Plasma | 0·8 (0·66–0·94) |
| P-tau231 | Alamar NULISA multiplex | CSF | 1 (1–1) |
| P-tau231 | MSD S-Plex | Plasma | 0·82 (0·68–0·96) |
| P-tau231 | MSD S-Plex | CSF | 1 (0·99–1) |
| P-tau231 | UGOT IPMS | Plasma | 0·96 (0·91–1) |
| P-tau231 | UGOT IPMS | CSF | N/A |
| P-tau231 | UGOT Simoa | Plasma | 0·82 (0·69–0·95) |
| P-tau231 | UGOT Simoa | CSF | 1 (1–1) |

**References**

1.	Lifke, V., Kollmorgen, G., Manuilova, E., Oelschlaegel, T., Hillringhaus, L., Widmann, M., von Arnim, C.A.F., Otto, M., Christenson, R.H., Powers, J.L., et al. (2019). Elecsys((R)) Total-Tau and Phospho-Tau (181P) CSF assays: Analytical performance of the novel, fully automated immunoassays for quantification of tau proteins in human cerebrospinal fluid. Clin Biochem *72*, 30-38. 10.1016/j.clinbiochem.2019.05.005.
