## Supplementary material for "The Alzheimer’s Association Global Biomarker Standardization Consortium (GBSC) plasma phospho-tau Round Robin study": STROBE statement

STROBE Statement—Checklist of items that should be included in reports of ***case-control studies***

|  | Item No | Recommendation | Manuscript section, page(s) | Comments |
| --- | --- | --- | --- | --- |
| **Title and abstract** | 1 | (*a*) Indicate the study’s design with a commonly used term in the title or the abstract | Abstract  p3 | Abstract background section: “case-control study” |
|  |  | (*b*) Provide in the abstract an informative and balanced summary of what was done and what was found | Abstract  p3 |  |
| Introduction | | |  |  |
| Background/rationale | 2 | Explain the scientific background and rationale for the investigation being reported | Introduction p7;  Research in Context panel p4-5 |  |
| Objectives | 3 | State specific objectives, including any prespecified hypotheses | Introduction p7 | “Our main aim was to compare all assays …” |
| Methods | | |  |  |
| Study design | 4 | Present key elements of study design early in the paper | Methods: participants, ethics, and study design p7 |  |
| Setting | 5 | Describe the setting, locations, and relevant dates, including periods of recruitment, exposure, follow-up, and data collection | Methods: participants, ethics, and study design subsection p7 | “Participant samples were selected serially …” |
| Participants | 6 | (*a*) Give the eligibility criteria, and the sources and methods of case ascertainment and control selection. Give the rationale for the choice of cases and controls | Methods: participants, ethics, and study design subsection p7 | “Participant samples were selected serially …” |
|  |  | (*b*) For matched studies, give matching criteria and the number of controls per case |  | N/A |
| Variables | 7 | Clearly define all outcomes, exposures, predictors, potential confounders, and effect modifiers. Give diagnostic criteria, if applicable | Methods: phosphorylated tau assays p8 |  |
| Data sources/ measurement | 8* | For each variable of interest, give sources of data and details of methods of assessment (measurement). Describe comparability of assessment methods if there is more than one group | Methods: phosphorylated tau assays p8; Supplementary methods |  |
| Bias | 9 | Describe any efforts to address potential sources of bias | Methods: participants, ethics, and study design subsection p7 | “blinded to participant information” |
| Study size | 10 | Explain how the study size was arrived at | Methods: participants, ethics, and study design subsection p7 |  |
| Quantitative variables | 11 | Explain how quantitative variables were handled in the analyses. If applicable, describe which groupings were chosen and why | Methods: statistical analysis p9 |  |
| Statistical methods | 12 | (*a*) Describe all statistical methods, including those used to control for confounding | Methods: statistical analysis p9 |  |
|  |  | (*b*) Describe any methods used to examine subgroups and interactions | Methods: statistical analysis p10 | “…we evaluated cross-matrix associations with Spearman correlation, calculated both in all patients and in the AD group” |
|  |  | (*c*) Explain how missing data were addressed | Methods: statistical analysis p10 |  |
|  |  | (*d*) If applicable, explain how matching of cases and controls was addressed | N/A |  |
|  |  | (*e*) Describe any sensitivity analyses | N/A |  |
| Results | | |  |  |
| Participants | 13* | (a) Report numbers of individuals at each stage of study—eg numbers potentially eligible, examined for eligibility, confirmed eligible, included in the study, completing follow-up, and analysed | Results: participant characteristics p10 |  |
|  |  | (b) Give reasons for non-participation at each stage | N/A |  |
|  |  | (c) Consider use of a flow diagram |  | Considered -not likely to add to understanding the text description |
| Descriptive data | 14* | (a) Give characteristics of study participants (eg demographic, clinical, social) and information on exposures and potential confounders | Results: Participant characteristics p10 and Table 2 |  |
|  |  | (b) Indicate number of participants with missing data for each variable of interest | Results: Groupwise differences of plasma and CSF p-tau assays p10; Candidate certified reference materials p12 |  |
| Outcome data | 15* | Report numbers in each exposure category, or summary measures of exposure | Results: Participant characteristics p10 and Table 2 |  |
| Main results | 16 | (*a*) Give unadjusted estimates and, if applicable, confounder-adjusted estimates and their precision (eg, 95% confidence interval). Make clear which confounders were adjusted for and why they were included |  | No confounders considered |
|  |  | (*b*) Report category boundaries when continuous variables were categorized | Methods: participants, ethics, and study design p8 | “A participant was considered to have “AD pathology” if the CSF results were Aβ42/Aβ40 <0·065 and p-tau181 >57 pg/mL.” |
|  |  | (*c*) If relevant, consider translating estimates of relative risk into absolute risk for a meaningful time period | N/A |  |

| Other analyses | 17 | Report other analyses done—eg analyses of subgroups and interactions, and sensitivity analyses | Supplementary figure 4  Supplementary figure 5 | Mean fold change  Areas under the ROC curves |
| --- | --- | --- | --- | --- |
| Discussion | | |  |  |
| Key results | 18 | Summarise key results with reference to study objectives | Research in cotext panel: Added value of this study p5 |  |
| Limitations | 19 | Discuss limitations of the study, taking into account sources of potential bias or imprecision. Discuss both direction and magnitude of any potential bias | Discussion p14-p15 |  |
| Interpretation | 20 | Give a cautious overall interpretation of results considering objectives, limitations, multiplicity of analyses, results from similar studies, and other relevant evidence | Research in context panel: Implications of all the available evidence p5 |  |
| Generalisability | 21 | Discuss the generalisability (external validity) of the study results | Discussion p14 |  |
| Other information | | |  |  |
| Funding | 22 | Give the source of funding and the role of the funders for the present study and, if applicable, for the original study on which the present article is based | Methods; role of the funding source |  |

*Give information separately for cases and controls.

**Note:** An Explanation and Elaboration article discusses each checklist item and gives methodological background and published examples of transparent reporting. The STROBE checklist is best used in conjunction with this article (freely available on the Web sites of PLoS Medicine at http://www.plosmedicine.org/, Annals of Internal Medicine at http://www.annals.org/, and Epidemiology at http://www.epidem.com/). Information on the STROBE Initiative is available at http://www.strobe-statement.org.
